# Mutation burden-orthogonal tumor genomic subtypes delineate responses to immune checkpoint therapy

**DOI:** 10.1101/2021.10.03.21264330

**Authors:** Shiro Takamatsu, Junzo Hamanishi, J.B. Brown, Ken Yamaguchi, Koji Yamanoi, Kosuke Murakami, Osamu Gotoh, Seiichi Mori, Masaki Mandai, Noriomi Matsumura

## Abstract

Higher-resolution tumor-agnostic biomarkers that predict response to immune checkpoint inhibitor (ICI) therapy are needed. Mutation signatures reflect underlying oncogenic processes that can affect tumor immunogenicity, and thus potentially delineate ICI treatment response among tumor types. Systematic mutational signature analysis on all solid tumors in The Cancer Genome Atlas yielded eight distinct tumor genomic subtypes, which were characterized by smoking exposure, ultraviolet light exposure, APOBEC enzyme activity, POLE mutation, mismatch repair deficiency, homologous recombination deficiency, genomic stability, and aging. The former five subtypes were construed to form an immune-responsive group acting as candidates for ICI therapy because of their high expression of immune-related genes and enrichment in cancer types with FDA approval for ICI monotherapy. Under this hypothesis, we developed a software to classify new tumors submitted to whole-exome sequencing. A pan-cancer dataset containing 973 cases treated with ICIs and clinical response was used for validation, where the classified tumor subtypes were significantly associated with ICI response, independent of cancer type and tumor mutational burden high or low status. The new tumor subtyping method can serve as a tumor-agnostic biomarker for ICI response prediction and will improve decision making in cancer treatment.

**Structured abstract:** *Background:* In cancer therapy, higher-resolution tumor-agnostic biomarkers that predict response to immune checkpoint inhibitor (ICI) therapy are needed. Mutation signatures reflect underlying oncogenic processes that can affect tumor immunogenicity, and thus potentially delineate ICI treatment response among tumor types.

*Methods:* Based on mutational signature analysis, we developed a stratification for all solid tumors in The Cancer Genome Atlas (TCGA). Subsequently, we developed a new software (GS-PRACTICE) to classify new tumors submitted to whole-exome sequencing. Using existing data from 973 pan-cancer ICI-treated cases with outcomes, we evaluated the subtype-response predictive performance.

*Results:* Systematic analysis on TCGA samples identified eight tumor genomic subtypes, which were characterized by features represented by smoking exposure, ultraviolet light exposure, APOBEC enzyme activity, *POLE* mutation, mismatch repair deficiency, homologous recombination deficiency, genomic stability, and aging. The former five subtypes were presumed to form an immune-responsive group acting as candidates for ICI therapy because of their high expression of immune-related genes and enrichment in cancer types with FDA approval for ICI monotherapy. In the validation cohort, the samples assigned by GS-PRACTICE to the immune-reactive subtypes were significantly associated with ICI response independent of cancer type and TMB high or low status.

*Conclusions:* The new tumor subtyping method can serve as a tumor-agnostic biomarker for ICI response prediction and will improve decision making in cancer treatment.

*One Sentence Summary:* A novel tumor genome subtyping method based on mutation signature analysis can predict response to immune checkpoint therapy orthogonally to tumor mutational burden.

## INTRODUCTION

The advent of immune checkpoint inhibitors (ICIs) has provided substantial opportunities in cancer treatment. However, the proportion of patients who benefit from ICIs varies widely by cancer type ^1^, and tumor-agnostic biomarkers to identify (un)responsive subsets are strongly desired. A recently established predictive biomarker is the loss of mismatch repair protein in immunohistochemistry or microsatellite instability (MSI-high), which indicates mismatch repair deficiency (MMRd) status ^2^. MMRd tumors are considered to be highly sensitive to ICI because they carry a large number of tumor-specific neoantigens ^3^. Yet another recently FDA-approved tumor agnostic biomarker is tumor mutational burden (TMB)-high status, where tumors have 10 or more mutations per megabase calculated from the FoundationOne CDx assay ^4^. Nevertheless, there are certainly cases that have modest TMB but respond to ICI ^5,6^, and more sophisticated methods for identifying such tumors need to be developed.

Comprehensive gene mutation analysis in cancer enabled by high-throughput next-generation sequencing has revealed that even neutral somatic mutations, previously thought to be “passenger” mutations, exhibit reproducible patterns of change, or mutational signatures, depending on the underlying endogenous and exogenous mutagenic processes ^7,8^. Certain mutational signatures are known to be associated with tumor immunogenicity ^9-11^, suggesting that differences in the background mutational processes may play an important role in anti-tumor immunity.

To advance oncology patient care by leveraging the signature-immunogenicity relationship, we report the development of a computational framework to classify tumors beyond their tissue origin. The tool is subsequently challenged to predict response to ICI independent of cancer type and TMB status using a large external patient dataset, demonstrating its feasibility and position to complement FDA-approved TMB analyses.

## RESULTS

### Identification of eight genomic subtypes based on mutational signature analysis

Based on Mutect2-derived ^12^ mutation annotations from whole-exome sequencing (WES) data, score profiles of COSMIC (v2) mutational signatures were derived for each solid tumor in The Cancer Genome Atlas (TCGA, n=9794). Eight tumor groups were obtained after clustering logarithm-transformed profiles (Figure 1A). Based on the enrichment of signatures with proposed etiologies (Table S1), seven of these subtypes were labeled as groups associated with smoking (SMK), ultraviolet light (UVL), APOBEC (APB), DNA polymerase epsilon deficiency (POL), mismatch repair deficiency (MRD), homologous recombination deficiency (HRD), and aging (AGE). The remaining group that showed no specific accumulation of mutation signatures and the lowest number of mutations was assigned the genomic stability (GNS) subtype.

**Figure 1.**
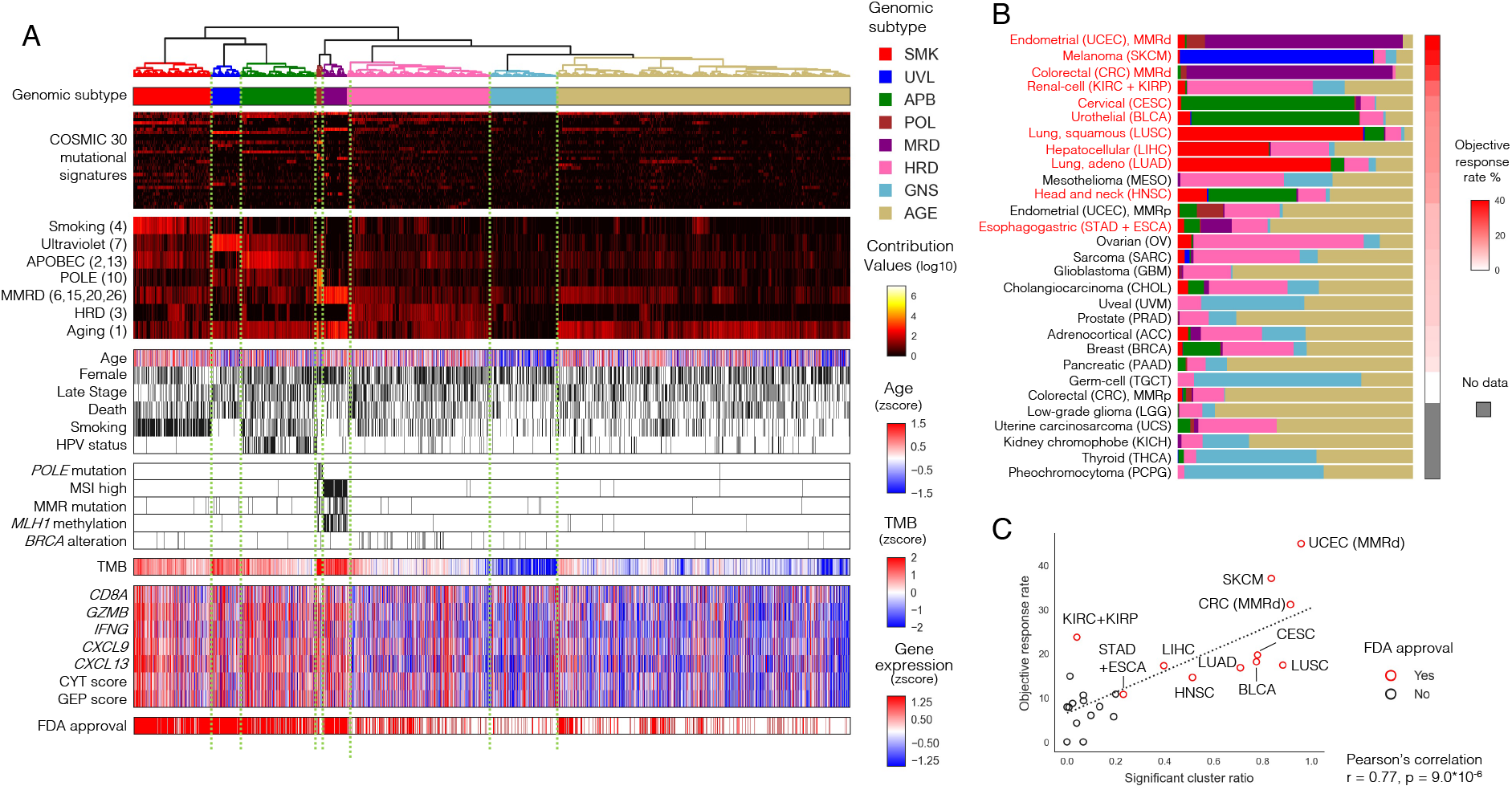
Identification of tumor genomic subtypes associated with immune responses. A) Unsupervised hierarchical clustering based on mutational signatures divided TCGA solid tumors (N=9794) into eight distinctive subtypes, five of which showed strong tumor immune responses. The first panel shows the genomic subtypes in color. The second panel shows COSMIC 30 mutational signature contribution values, and the third panel highlights the seven known etiology-related ones, with COSMIC mutational signature numbers in parentheses. The fourth panel shows clinical information (age, sex, stage, death, smoking habits, HPV infection), and the fifth panel shows DNA repair-related gene alterations, including *POLE* mutations, MSI high, MMR mutations, *MLH1* methylation, and *BRCA* alterations are shown in this order. The sixth panel shows TMB, and the seventh panel shows immune-related gene expression (*CD8A, GZMB, IFNG, CXCL9*, and *CXCL13*) and scores (CYT and GEP). The bottom panel shows the cancer types for which ICI therapy as a single agent is approved by the FDA. These results are summarized in Figure S1A and Figure S1B. B) The distribution of the subtypes (shown with the same colors as A) and reported objective response rate to ICI monotherapy per tumor type. Tumor types are arranged in descending order of the response rate, with red letters indicating those with FDA approval for ICI monotherapy. C) The proportion of the five subtypes with high immune-responsive scores (SMK, UVL, APB, POL, and MRD) per tumor type showed a significant positive correlation with the reported objective response rate to ICI monotherapy for that type. MMRd; mismatch repair deficiency, MMRp; mismatch repair proficiency

In terms of clinical information, age, gender, disease stage, and mortality differed considerably among the subtypes (Figure 1A, Figure S1A). The proportion of patients with smoking history was highest in the SMK group. Molecularly characterized groups also contained enriched annotations, including high *POLE* mutations in the POL group, as well as MMR mutations, *MLH1* methylation, and MSI high status in the MRD group. The HRD group contained characteristic *BRCA* alterations ^13^. A detailed stratification of subtypes per primary tumor origin reveals distinct partitions per type (Figure 1B, S1B, S1C). Extensive analytics of each subtype are provided in Figure S1D and S2-S8.

Transcriptomes of genes associated with tumor immune response were assessed. Genes representing the infiltration of cytotoxic CD8+ T cells (*CD8A, GZMB*, and *IFNG*) and genes related to ICI response (*CXCL9* and *CXCL13*) ^14^ were upregulated in the five subtypes (SMK, UVL, APB, POL, MRD) relative to the others (HRD, GNS, AGE). The CYT score ^15^ and GEP score ^16^ related to ICI response were also higher in the same five subtypes. Post-subtyping also demonstrates that the five subtypes were more frequently of tumor origin with FDA approval for ICI monotherapy (Figure 1A). Further, when the proportion of samples assigned to the five subtypes was scored per tumor type, the score was strongly correlated with the previously reported objective response rate to ICI monotherapy for that tumor type (Figure 1C) ^1,17,18^. The SMK/UVL/APB/POL/MRD subtypes thus serve to prognosticate positive response to ICI administration, and are hereafter termed immuno-responsive genomic subtypes (irGS).

### Development of GS-PRACTICE

A software tool embedding machine learning was developed to stratify newly sequenced tumors into the eight genomic subtypes derived above (Figure 2A). First, hierarchical clusters were again derived using each of three alternative variant calling schemes (Figure S9A, see Methods). High concurrence with analyses based on Mutect2 was observed (Figure S9A, S9B). To extract samples typical for each subtype as a training dataset, samples with matching classification results in at least three of the four methods, including concomitant classification with Mutect2, were selected and used for subsequent analysis (Figure S9B, S9C). The resulting 7181 samples and their 30 COSMIC signature scores were used as features to construct Nearest Neighbor, Support Vector Machine, Random Forest, and Logistic Regression classifiers with optimized hyperparameters (see Methods and Figure S10A). All classifiers showed more than 95% subset accuracy (exact match ratio) in multi-label classification (Figure S10B), yielding a robust 8-class ensemble-based stratification tool.

**Figure 2.**
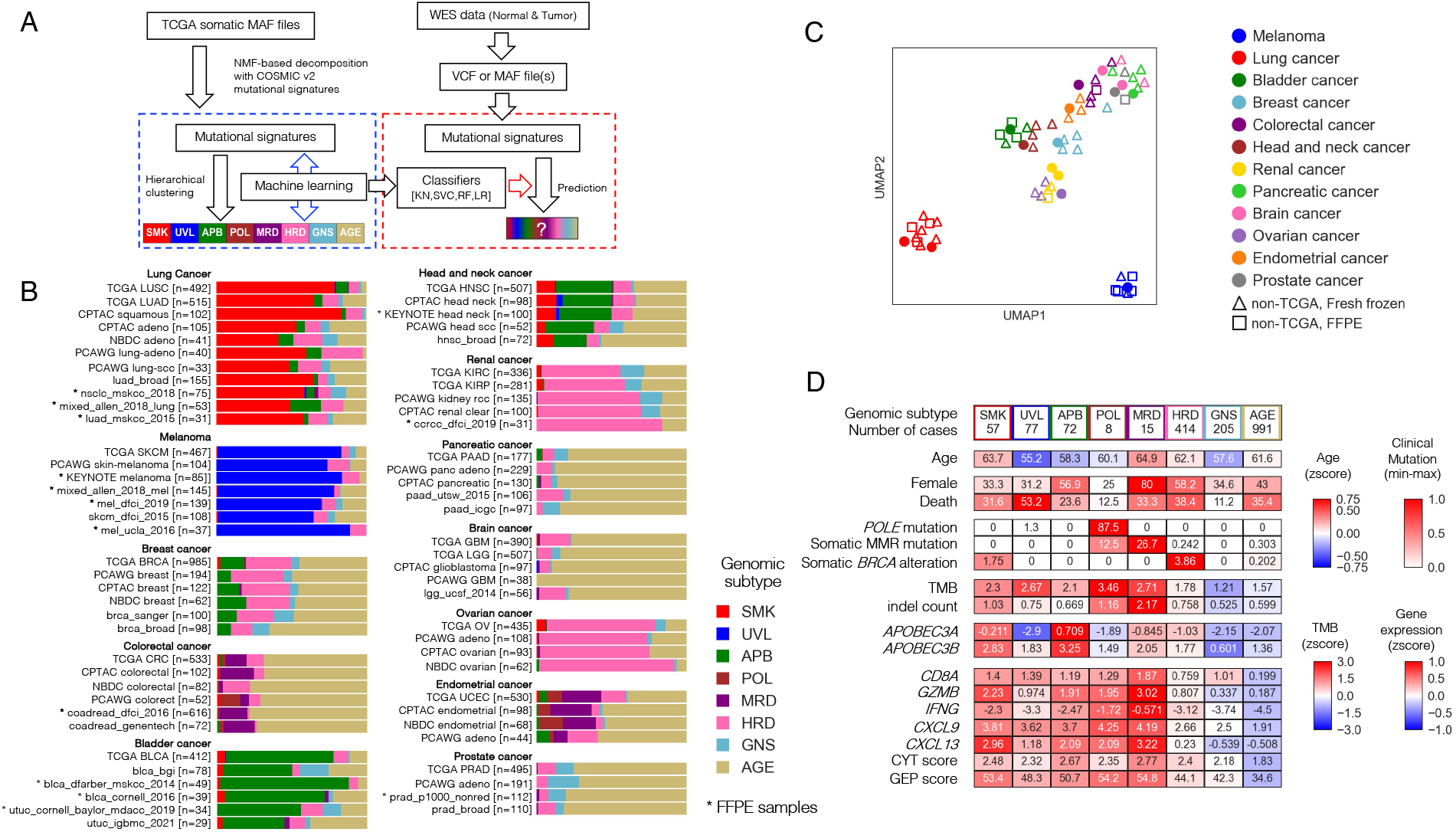
Development of GS-PRACTICE. A) Overview of the program. Using the TCGA dataset, four different classifiers were built from four different algorithms, namely k-nearest neighbor (KN), support vector machine (SVC), random forest (RF), and logistic regression (LR). Using external somatic mutation profiles from WES data as input, the four classifiers output classification results. B) Subtyping results by GS-PRACTICE for each cancer type in the publicly available data (details in Table S2). Asterisks indicate data obtained from FFPE samples, which are similar to data obtained from frozen samples. Note that for NBDC colorectal cancer, the percentage of MMRd tumors has been reported to be low in Japanese^49^. C) UMAP plot using the proportion of assigned subtypes as feature values leading to spatial projection. Marker color indicates the derived organ. Dot markers indicate TCGA data, triangles indicate non-TCGA data from frozen samples, and squares indicate non-TCGA data from FFPE samples. Datasets with the same cancer type are adjacent to each other, indicating a similar distribution of genomic subtypes across differing data sources. D) Comparison between the genomic subtypes in PCAWG datasets with multiple cancer types (n=1916). Immune-related gene expression and scores were higher in irGS. The distribution of genomic subtypes in individual cancer types are indicated in Figure 2B and Figure S11B. GS-PRACTICE; Genomic Subtyping and Predictive Response Analysis for Cancer Tumor ICi Efficacy, CPTAC; Clinical Proteomic Tumor Analysis Consortium, NBDC; National Bioscience Database Center, MMRd; mismatch repair deficiency, PCAWG; Pan-Cancer Analysis of Whole Genomes consortium, irGS, immune-reactive genomic subtype

For new query inputs of somatic mutation profile scores derived from tumor sequencing, each of the four classifiers is executed, and predictions are deemed consistent when the three or four resultant classifications concur; otherwise a classification of undeterminable (UND) is assigned to the sample. Further, when a majority prediction is one of the SMK, UVL, APB, POL, or MRD subtypes, the subtype is additionally classified as irGS; otherwise it is labeled as non-irGS. The prediction system, GS-PRACTICE (acronym for “Genomic Subtyping and Predictive Response Analysis for Cancer Tumor ICi Efficacy”), is publicly available in the GitHub page (https://github.com/shirotak/GS-PRACTICE).

GS-PRACTICE was tested for its ability to stratify a diverse collection of samples from various sources into genomic subtypes (Table S2). Collectively, 96-98% of samples were successfully assigned a subtype, indicating consensus in the ensemble’s individual classifiers. The classifier concordance rate was also consistent across different data sources, and was consistent irrespective of whether samples were of FFPE or frozen tissue origin (Figure S10C). Re-analyses restricted to individual cancer types yielded identical conclusions with respect to tissue origin and source (Figure 2B, 2C).

We applied GS-PRACTICE to 1916 samples of the Pan-Cancer Analysis of Whole Genomes consortium (PCAWG) datasets ^19^ using somatic mutation profiles in coding regions obtained from the UCSC Xena ^20^ as input (Figure S11A, S11B). Results paralleled that of TCGA data (Figure 2D). The differences in age, sex, and mortality among the subtypes were similar to those of TCGA. Somatic *POLE* mutations were common in the POL group, somatic MMR mutations in the MRD group, and somatic *BRCA* alterations (*BRCA1/2* mutations with LOH) in the HRD group. APOBEC3 family gene expression was elevated in the APB group. The five irGS subgroups demonstrated increased gene expression and biomarker scores associated with infiltration of cytotoxic CD8+ T cells and ICI response (Figure 2D, Figure S1A, S4A).

### GS-PRACTICE as a tumor agnostic predictive biomarker for ICI response

973 cases with information on objective response to ICI treatment were used to challenge and assess the subtyping and (non-)irGS assignment from GS-PRACTICE (Table S3). Taken in total, ICI response rate was significantly higher in irGS than non-irGS (34.6% vs 12.0%, P = 5.1 ×10^−14^, Figure 3A). When analyzed by the eight subtypes, the five subtypes belonging to irGS tended to have a higher response rate than the three non-irGS subtypes (Figure S12).

**Figure 3.**
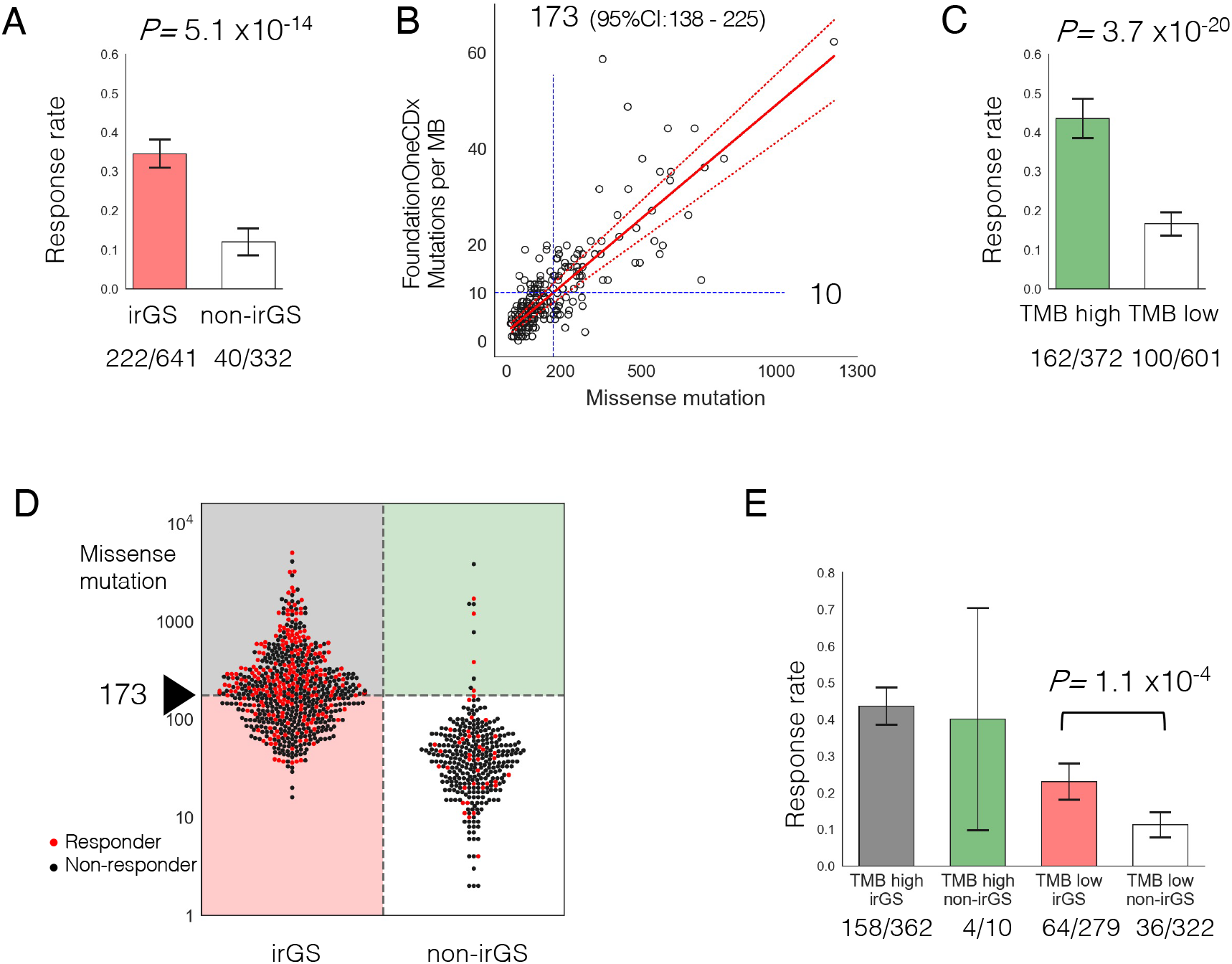
Prediction of ICI response by GS-PRACTICE and TMB. A) ICI response rate was significantly higher in irGS tumors than non-irGS. B) Determination of TMB cutoff. The bladder cancer dataset (n=218) from Mariathasan et al. ^22^ was examined. The number of missense mutations in the whole-exome sequencing and the number of mutations per megabase from the FoundationOneCDx panel assay were plotted. From the regression equation, 10 mutations per megabase in the panel corresponded to 173 missense mutations (95% confidence interval:138 - 225). C) ICI response rate was significantly higher in TMB high tumors than TMB low tumors. D) Association between distribution of TMB and ICI response per sample divided by irGS status. Red dots indicate responders, and black dots indicate non-responders. E) Comparison of ICI response rates in the four groups stratified by irGS and TMB status. irGS tumors had a significantly higher response rate than non-irGS within the samples classified as TMB low. ICI, immune checkpoint inhibitor; irGS, immune-reactive genomic subtype; TMB, Tumor mutational burden

Next, to determine a cutoff for assignment of TMB-high, we compared the number of mutations detected in our WES pipeline with those in FoundationOne CDx using a bladder cancer dataset ^22^. Based on Passing-Bablok regression analysis, the cutoff of 10 mutations per megabase in the CDx panel corresponds to 173 missense mutations in a WES sample (95% confidence interval of 138-225) (Figure 3B). Using this value as the cutoff for TMB-high, tumors categorized as TMB-high showed higher ICI response rate than those as TMB-low (43.5% vs 16.6%, P = 3.7 ×10^−20^, Figure 3C).

When we divided the tumors into four groups according to the pairwise stratifications of (non-)irGS and TMB-low/high, 97.2% of TMB-high tumors belonged to irGS and 96.9% of non-irGS tumors belonged to TMB-low (Figure 3D). Response rate to ICI was highest in the TMB-high irGS group (43.6%). Critically, within TMB-low tumors, irGS tumors had a significantly higher response rate than non-irGS (22.9% vs 11.2 %, *P=* 1.1 ⨯10^−4^, Figure 3E). Additionally, in a multivariate logistic regression analysis, irGS status was significantly associated with the objective response to ICI after adjustment for TMB-high status and cancer type (adjusted odds ratio, 2.18; 95% confidence interval, 1.40-3.40; *P=* 5.6 ⨯10^−4^, Figure 4). The trends were similar when examined separately by anti-PD-1 antibody or anti-PD-L1 antibody therapy, as well as by anti-CTLA-4 monotherapy and anti-CTLA-4/anti-PD-1 combination therapy (Figure S13). These results were also significant when limited to data from the KEYNOTE clinical trials (n=311), a prospective cohort of patients treated solely with anti-PD-1 antibody, pembrolizumab (Figure S14). Although the KEYNOTE trials excluded patients with clinically diagnosed MMRd tumors at enrollment, two tumors from the cohort (one each with gastric cancer and biliary tract cancer) were classified into the MRD subtype, and both of them responded to ICI. Furthermore, the results were similar when using the cohort’s optimal TMB cutoff determined by the ROC curve and the Youden index or using log(10)-transformed TMB as a continuous variable (Figure S15). Even when the recently reported score for estimating T-cell infiltration in tumors from WES data ^21^ was added as a covariate, there remained a significant correlation between irGS and ICI response (Figure S16). Genome subtyping and ICI response analysis by GS-PRACTICE for each of the 13 individual ICI studies comprising the combined 973 patients are described in Figure S17 and Table S4.

**Figure 4.**
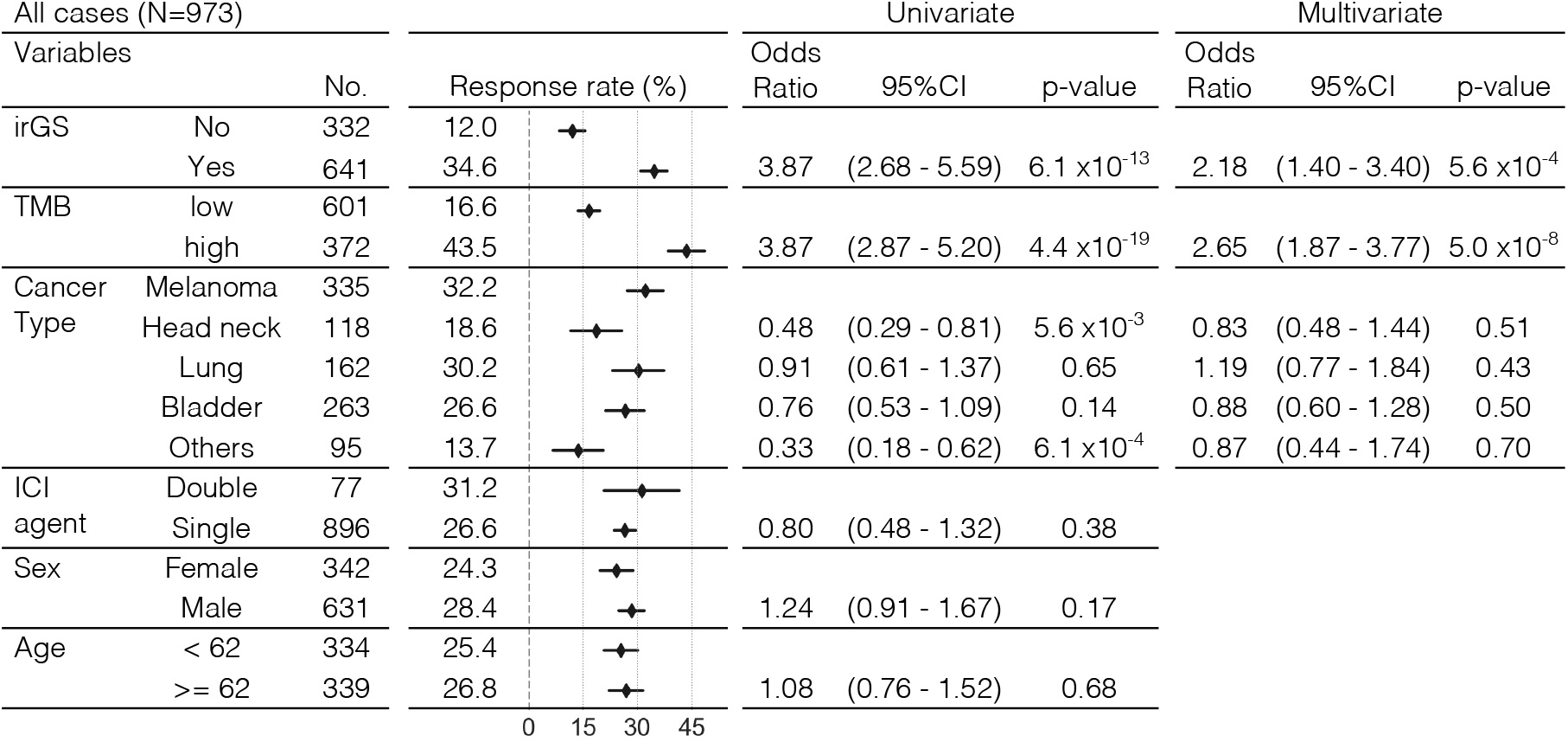
Univariate and multivariate logistic regression analysis for ICI response in the validation cohort. Multivariate logistic regression analysis showed that irGS was significantly associated with the ICI response after adjusting by TMB status and cancer type. irGS, immune-reactive genomic subtype; TMB, Tumor mutational burden; Single, either anti-PD-1, anti-PD-L1, or anti-CTLA4 antibody; Double, combination of anti-CTLA4 and anti-PD-1 or anti-PD-L1 antibodies; OR, Odds ratio; CI, confidence interval

## DISCUSSION

The relationship between mutational signatures and ICI response has been previously reported for several specific types of cancer. For example, mutational signatures in melanoma ^10^ and non-small cell lung cancer ^11^ correlate with response to ICI, and these data are explained by the idea that the process of carcinogenesis by exogenous mutagens (UV, tobacco) results in highly immunogenic tumor antigens ^23^. In addition, APOBEC-related mutational signatures are associated with viral infections and a specific mutational pattern called kataegis, which also produces highly immunogenic antigens ^24,25^ and is associated with ICI response in non-small cell lung cancer ^26,27^. On the other hand, it has been reported that high copy number, aneuploidy, and HRD-associated scores inversely correlate with tumor immune response ^28-30^, and negative results in a recent clinical trial in ovarian cancer where half of the tumors showed HRD ^31,32^ suggest that HRD-related signatures are unlikely to be associated with high sensitivity to ICIs. Aging-related (clock-like) mutational signatures are reported to be associated with lower immune activity in melanoma and non-small cell lung cancer treated with ICI ^9,33^. Since many age-related gene mutations also occur in non-tumor cells ^34^, they may be related to immune tolerance. Our categorization of irGS and non-irGS in this study is supported by previous reports on the relationship between specific mutation signatures and tumor immunogenicity, and provides a cross-organ assessment of this relationship.

In June 2020, the FDA approved pembrolizumab for the treatment of tumors diagnosed 10 mutations per megabase or greater by FoundationOne CDx ^4^. This cutoff corresponded to 173 missense mutations in our WES analysis (Figure 3B) and was close to the optimal cutoff value of 165 calculated by the ROC curve based on the collated cohort we assembled (Figure S14). However, TMB quantification based on the panel assay is still subject to fluctuation (Figure 3B), which may impact clinical decisions, and thus, comprehensive sequencing methods including WES are considered optimal for reproducible and reliable measurements ^35^. As the cost of WES decreases and efforts toward the implementation of WES as a routine cancer treatment continue to advance ^36,37^, the combination of precision-improved TMB calculation and the orthogonal GS-PRACTICE method will usher in precise patient selection for ICI treatment.

There have been some criticisms that there is no logical basis for setting a universal TMB threshold for all solid tumors, since such an index is a continuous value that varies considerably among cancer types ^7,38,39^. Our analysis showed that almost all non-irGS tumors belonged to TMB-low (Figure 3D, S13, S14, and S15), indicating that the current TMB cutoff has the consequence to exclude non-irGS tumors, which have no or little immunogenic background mutational processes. In other words, our method may add biological rationales to the empirically determined TMB cutoff.Additionally, the previous report that the optimal cutoffs for TMB-high differed among cancer types ^5^ may be explained by the different distribution of genomic subtypes per tumor origin (Figure 1B).

While GS-PRACTICE represents an advance in cancer diagnostics and clinical decision making, some limitations of this work must be made transparent. First, due to lack of data, prolonged survival or improved quality of life in ICI-responsible irGS patients could be quantitatively validated. For this purpose, design and logistics of appropriate randomized control trials using ICI are needed. Secondly, accurate subtyping may not be possible for tumors with a small number of mutations due to computational reasons. The clustering results using the four variant callers showed relatively low concordance rates for the HRD, AGE, and GNS subtypes (Figure S9B). Renal cancers had a moderately low number of mutations and were mostly classified as HRD, but their HRD scores and indel signature 6 ratios were low (Figure S7B), indicating that they are unlikely to have homologous recombination deficiency properties. It is known that the response to ICI in renal cancer is not associated with TMB ^40^, and the present analysis also did not identify any characteristic mutation patterns associated with ICI response. One method to improve on the state of the art would be to apply GS-PRACTICE to whole genome sequencing, which can detect dozens of times more mutations than WES ^8^. This may allow for higher resolution mutation signature analysis and more sophisticated tumor genome subtyping even in tumors with a small number of coding mutations.

GS-PRACTICE represents a pan-cancer advancement in both solid tumor diagnostics and precision medicine, as it subtypes tumors by leveraging mutational signatures with defined etiologies, and the subtypes were shown to be indicative of ICI response. The method can be reproducibly applied to WES data derived from FFPE specimens, and thus immediately provide a predictive biomarker for ICI treatment in clinical practice. Future analyses of randomized control trials and whole genome sequencing will spur improved dataset generation for model building, which will subsequently strengthen the clinical utility of the protocol developed herein.

## METHODS

### TCGA data

Clinical information of all tumors except diffuse large B-cell lymphoma, acute myeloid leukemia, and thymoma in TCGA studies was obtained from the cBioPortal (https://www.cbioportal.org/) and the broad GDAC websites (https://gdac.broadinstitute.org/). Among these, 9794 cases, whose somatic mutation profiles analyzed by Mutect2 ^12^ were available on the GDC portal (https://portal.gdc.cancer.gov/), were included in this study. We also obtained the other somatic mutation profiles calculated by the three different variant callers (Varscan2 ^41^, MuSE ^42^, Somatic Sniper ^43^) and gene expression profiles from a previous report ^44^. The annotations of germline mutations and gene promoter methylations were obtained from previous reports ^45, 46^. The contribution values to COSMIC (v2) 30 mutational signatures (https://cancer.sanger.ac.uk/signatures/signatures_v2) of each sample were calculated using MutationalPatterns ^47^. The annotation of cancer types with FDA approval for ICI monotherapy was based on a previous report ^48^. The response rates for ICI monotherapy for each tumor type were obtained from previous reports ^1, 17 18,^

### Validation datasets

PCAWG, CPTAC, NBDC, and cBioPortal datasets were obtained from their databases (Table S2). For the ICI-treated cohorts, samples collected from metastatic tumors and those with a history of ICI treatment at sample collection were excluded. A total of 973 patients from 13 datasets were included in the analysis (Table S3, Figure S15).

### Statistical analyses

Statistical analyses were mainly performed in Python (3.7.4); the Mann–Whitney U test, chi-square test, and Spearman’s rank correlation coefficient test were performed using SciPy (1.6.1), survival analyses including the Kaplan–Meier curve, log-rank test, and Cox proportional hazard regression using Lifelines (0.25.10) and StatsModels (0.12.2), machine learning analyses using Scikit-learn (0.24.1). The Venn diagram, the Jonckheere-Terpstra test, and the Passing-Bablok regression analysis were performed using “VennDiagram” (1.6.20), “clinfun” (1.0.15), and “mcr” (1.2.2) packages in R. We considered a p-value < 0.05 as being statistically significant.

## Data Availability

Controlled access data used in this study were obtained from dbGaP, EGA, and NBDC with access permissions according to the respective required procedures. The processed data and analysis codes to reproduce the results of this work are available on the GitHub page (https://github.com/shirotak/pancancer_MutSig_ICI). Other codes for preprocessing or restricted-access data are available from the corresponding author upon reasonable request.

## Supplementary data

Supplementary methods

Figure S1. Characteristics of the eight tumor genomic subtypes derived from TCGA solid tumors (N=9794)

Figure S2. Features of SMOKING (SMK) subtype (n=1072)

Figure S3. Features of ULTRAVIOLET LIGHT (UVL) subtype (n=401)

Figure S4. Features of APOBEC (APB) subtype (n=1036)

Figure S5. Features of MISMATCH REPAIR DEFICIENCY (MRD) subtype (n=339)

Figure S6. Features of *POLE* (POL) subtype (n=81)

Figure S7. Features of HOMOLOGOUS RCOMBINATION DEFICIENCY (HRD) subtype (n=1956)

Figure S8. Features of GENOMICALLY STABLE (GNS) subtype (n=909)

Figure S9. Consistency of clustering results from different variant callers

Figure S10. Developing classifiers through machine learning algorithms

Figure S11. Tumor subtyping by GS-PRACTICE in the PCAWG datasets

Figure S12. Relationship between tumor genomic subtype, response rate and cancer type in the whole cohort

Figure S13. Studies examined by type of drug

Figure S14. Study in the dataset from the KEYNOTE trials, which were prospective cohort studies of patients treated with solely pembrolizumab (n=311)

Figure S15. Studies using the cohorts’ optimal TMB cutoff or logarithmic TMB as a continuous value

Figure S16. Study using TcellExTRECT score

Figure S17. Association between TMB and ICI response divided by irGS status per dataset Figure S18. Determination of MSI-high cases using MSIsensor

Figure S19. Comparison of the number of missense mutations or non-synonymous SNVs from our WES pipeline and previously published data

Table S1. COSMIC version2 mutational signatures and proposed etiologies Table S2. Datasets for evaluating subtype classification

Table S3. Datasets for evaluating ICI treatment response

Table S4. Per-sample annotations of the ICI response validation cohort

## Acknowledgments

We acknowledge all the researchers and companies for providing the academic and industrial datasets used in this study. Some of these data were originally obtained by Japanese researchers and are available on the website of the National Bioscience Database Center (NBDC).

## Author contributions

Conceptualization: ST, JH, JBB, NM

Data curation: ST, JH

Formal analysis: ST, OG, KM

Funding acquisition: JH, NM

Investigation: ST, JH, KYamaguchi, KYamanoi

Methodology: ST, JBB, NM

Project administration: JH, MM

Resources: JH, SM, MM

Software: ST, JBB

Supervision: NM, SM, MM

Visualization: ST, OG

Writing – original draft: ST, JBB, NM

Writing – review & editing: ST, JH, JBB, KYamaguchi, KYamanoi, OG, KM, NM

## Data and code availability

Controlled access data used in this study were obtained from dbGaP, EGA, and NBDC with access permissions according to the respective required procedures (Table S2 and S3). The processed data and analysis codes to reproduce the results of this work are available on the GitHub page (https://github.com/shirotak/pancancer_MutSig_ICI). Other codes for preprocessing or restricted-access data are available from the corresponding author upon reasonable request.

