## Supplementary methods and figures for "Mutation burden-orthogonal tumor genomic subtypes delineate responses to immune checkpoint therapy"

### Supplementary data

#### Supplementary methods

**Analysis of TCGA samples**

**Analysis of non-TCGA datasets**

**Statistical analyses**

**Data and code availability**

#### Supplementary figures

**Figure S1. Characteristics of the eight tumor genomic subtypes derived from TCGA solid tumors (N=9794)**

**Figure S2. Features of SMOKING (SMK) subtype (n=1072)**

**Figure S3. Features of ULTRAVIOLET LIGHT (UVL) subtype (n=401)**

**Figure S4. Features of APOBEC (APB) subtype (n=1036)**

**Figure S5. Features of MISMATCH REPAIR DEFICIENCY (MRD) subtype (n=339)**

**Figure S6. Features of *POLE* (POL) subtype (n=81)**

**Figure S7. Features of HOMOLOGOUS RCOMBINATION DEFICIENCY (HRD) subtype (n=1956)**

**Figure S8. Features of GENOMICALLY STABLE (GNS) subtype (n=909)**

**Figure S9. Consistency of clustering results from different variant callers**

**Figure S10. Developing classifiers through machine learning algorithms**

**Figure S11. Tumor subtyping by GS-PRACTICE in the PCAWG datasets**

**Figure S12. Relationship between tumor genomic subtype, response rate and cancer type in the whole cohort**

**Figure S13. Studies examined by type of drug**

**Figure S14. Study in the dataset from the KEYNOTE trials, which were prospective cohort studies of patients treated with solely pembrolizumab (n=311)**

**Figure S15. Studies using the cohorts' optimal TMB cutoff or logarithmic TMB as a continuous value**

**Figure S16. Study using TcellExTRECT score**

**Figure S17. Association between TMB and ICI response divided by irGS status per dataset**

**Figure S18. Determination of MSI-high cases using MSIsensor**

**Figure S19. Comparison of the number of missense mutations or non-synonymous SNVs from our WES pipeline and previously published data**

#### Supplementary tables

**Table S1. COSMIC version2 mutational signatures and proposed etiologies**

**Table S2. Datasets for evaluating subtype classification**

**Table S3. Datasets for evaluating ICI treatment response**

**Table S4. Per-sample annotations of the ICI response validation cohort**

#### References

### Methods

#### Analysis of TCGA samples

##### Sample selection and collection of clinical information

Clinical and somatic gene mutation profiles of all tumors except diffuse large B-cell lymphoma, acute myeloid leukemia, and thymoma, PanCancer Atlas datasets were downloaded from cBioPortal [1] (29 studies, 10075 cases). Information on smoking habit and HPV infection status was obtained from GDAC [2]. Among these tumors, 9794 cases for which the somatic mutation profiles analyzed by Mutect2 [3] in the MAF format were available from the GDC portal [4] were selected for analysis.

The annotation of cancer types with FDA approval for ICI monotherapy was based on a previous report [5]. The response rates for ICI monotherapy for each tumor type were obtained from previous reports [6, 7]. The response rate data for endometrial cancer with mismatch repair deficiency or with mismatch repair proficiency were calculated from another previous report [8].

##### Identification of genomic subtypes based on mutational signatures

Using MutationalPatterns [9], the contribution values of each sample to COSMIC v2 30 mutational signatures [10] were calculate from the four deferent somatic mutation profiles, which were pre-computed using Mutect2 [3,] Varscan2 [11], MuSE [12], and Somatic Sniper [13] and were available in the MAF format on the GDC portal [4]. Using the log10 transformed values of these contribution values, unsupervised hierarchical clustering with Ward's method was performed.

##### Annotation of gene alterations

Somatic gene mutations annotated as significant in the PanCancer Atlas studies of cBioPortal [1] were included, while those marked as mutations of unknown significance were excluded. For germline mutations, we obtained the annotations from a previous report [14], where those annotated as “likely pathogenic” or “pathogenic” in “Overall Classification” column were retained. For gene mutations in *BRCA1* and *BRCA2*, we extracted those with locus-specific LOH or homozygous deletions as we previously reported [15]. For gene promoter methylations in *MLH1* and *BRCA1*, we obtained annotations from a previous report [16].

##### Insertions and deletions-based mutational signature 6

To investigate the features of HRD subtype tumors, we calculated indel signature 6, a signature derived from small insertions and deletions (ID) that has been reported to be associated with the HRD phenotype in recent whole genome sequencing studies [17]. The annotated somatic mutations called by Mutect2 were obtained from the GDC portal [4] in the VCF format, and the insertions and deletions with "PASS" annotations were extracted. The contribution values of each sample to the COSMIC reference ID signatures [10] were calculated using YAPSA [18]. The ratio of contribution values of indel signature 6 to the number of all detected mutations were calculated as indel signature 6 ratio.

##### MSI score and MSI-high annotation

We calculated MSI scores of all samples using MSI sensor [19] with the default parameters from normal-tumor paired whole exome sequencing (WES) data. For UCEC, CRC, STAD, and ESCA, MSI status was obtained from the clinical information in cBioPortal. Within these samples, the optimal cutoff value of MSI score for the annotated MSI-high cases was calculated using the ROC curve and the Youden index (Figure S18A,B). Then, for the other cancer types, samples with the score above this cutoff were determined to be MSI-high (Figure S18C).

##### Other genomic alterations scores

The following scores were calculated from the somatic mutation profiles calculated by Mutect2. Tumor mutational burden: the number of missense mutations. Total indel count: the total number of frameshift insertions, inframe insertions, frameshift deletions, and inframe deletions. Indel ratio: the ratio of the total indel count to the total number of detected mutations, including synonymous mutations. Insertion to indel ratio: the ratio of the total number of frameshift insertions and inframe insertions to the total indel count.

For predicted neoantigens counts based on netMHCpan [20], we obtained the pre-computated data for SNVs and indels from a previous report [21].

For chromosomal changes, we obtained HRD scores and CNV burden scores from the GDC portal [4].

**Gene expression scores**

We obtained batch-corrected gene expression values from a previous report [21]. The CYT score was calculated from the geometric mean of the expression levels of *GZMA* and *PRF1* according to the previous report [22]. We obtained a gene set "HALLMARK_PI3K_AKT_PATHWAY" from MSigDB [23] and calculated its enrichment score using ssGSEA [24]. In addition, using the literature [25] as a reference, the GEP score was calculated as the sum of each gene expression multiplied by the following coefficients: *CL5*=0.008346; *CD27*=0.072293; *CD274*=0.042853; *CD276*=-0.0239; *CD8A*=0.031021; *CMKLR1*=0.151253; *CXCL9*=0.074135; *CXCR6*=0.004313; *HLA.DQA1*=0.020091; *HLA.DRB1*=0.058806; *HLA.E*=0.07175; *IDO1*=0.060679; *LAG3*=0.123895; *NKG7*=0.075524; *PDCD1LG2*=0.003734; *PSMB10*=0.032999; *STAT1*=0.250229; *TIGIT*=0.084767.

**TcellExTRECT score**

BAM files from primary tumor samples of skin cutaneous melanoma (SKCM), head and neck squamous cell carcinoma (HNSC), lung adenocarcinoma (LUAD), lung squamous cell carcinoma (LUSC), and bladder urothelial carcinoma (BLCA) were downloaded from the GDC portal and used as input to calculate the TcellExTRECT score as previously reported [26]. Samples with 'qcFit' values greater than 4 were excluded from the analysis. Because adjustments for 'TCRA.fraction.score' by known tumor purity and copy number status did not improve the positive correlation with T-cell related gene expressions, the score was used without such adjustments.

**Development of GS-PRACTICE**

Using the 30 signature contribution values as features and the genomic subtypes as labels in the selected 7181 samples (Figure S9C), we built four independent classifiers from four different algorithms, namely, k-nearest neighbor, support vector machine, logistic regression, and random forest using the Scikit-learn module in Python. For the former three classifiers, the main hyperparameters were optimized by double cross-validation (Figure S10A). First, the above selected TCGA samples were divided into two parts, X1 and X2. Second, parameters were calculated using X1 by two-fold cross-validation, and those parameters were evaluated using X2 as test data. Third, X1 and X2 were swapped, and the same calculations were performed. These processes were repeated 100-1000 times to determine the optimal parameters (Figure S10A). For the random forest model, since parameter adjusting hardly changed the prediction accuracy, the default settings were used.

After calculating the contribution values of the 30 mutational signatures from external somatic mutation profiles in VCF or MAF format using MutationalPatterns [9], the four classifiers independently make predictions using these values as input, and then integrate the results to output the final classification (Figure 2A). In the classification into eight subtypes, if the results from three or more classifiers matched, the matched result was determined to be the subtype, otherwise it was determined to be undeterminable (UND). In parallel, if the results from three or more classifiers belong to immune-related genomic subtype (irGS), namely, SMK, UVL, APB, POL, or MRD, it was classified as irGS, otherwise non-irGS. This tool is available in the GitHub page (https://github.com/shirotak/GS-PRACTICE).

**Analysis of non-TCGA datasets**

**Pan-Cancer Analysis of Whole Genomes consortium (PCAWG) datasets [27]**

Donor-centric phenotype data, consensus coding mutation profiles, allele-specific copy number profiles, RNAseq-based gene expression profiles were obtained from the UCSC Xena [28]. From a total of 2834 cases, those included in the whitelist, with consensus coding mutations available, aged 20 years or older, and with tumor types included in TCGA studies were analyzed (Figure S11A). Subtyping by GS-PRACTICE was performed using the consensus coding mutations in the MAF format as input (Figure S11B). Somatic mutations were retained only when the altered allele count was ≥ 4, the altered allele frequency was ≥ 2.5%, and the oncoKB [29] annotation was either 'pathogenic' or 'likely pathogenic'. Somatic *BRCA1/2* mutations were checked for their allele-specific copy numbers, and only those with the minor allele copy number equal to zero were retained as significant alterations, as previously reported [15]. The normalized gene expression values, derived from the log2 transformed upper quartile fragments per kilobase of transcript per million mapped reads (FPKM-UQ), were used for analysis. The CYT score [22] and the GEP score [25] were calculated in the same way as TCGA data analysis.

**Clinical Proteomic Tumor Analysis Consortium (CPTAC) datasets [30]**

Clinical information and somatic mutation profiles in the MAF format were obtained from GDC portal [4].

**cBioPortal datasets [1]**

Clinical information and somatic mutation profiles in the MAF format were obtained from the websites (Table S2).

**National Bioscience Database Center (NBDC) datasets [31]**

We obtained clinical and raw WES data (Table S2) through the application process for NBDC Human Database [31], calculated somatic mutation profiles in our WES analysis pipeline (see below).

**Sample selection and definition of response in ICI-treated cohorts**

Collected samples were derived from pairs of the primary tumor and normal blood or tissue, and those collected from metastatic sites different from the primary tumor (lymph nodes, bones, distant internal organs, etc.) were excluded. In addition, collected samples were taken before or during ICI administration, and those with a history of ICI treatment at the time of sample collection were excluded. Most of the cases were evaluated using the RECIST criteria for radiological response or equivalent, where CR/PR was defined as a responder and SD/PD/NE as a non-responder. In some datasets, we could not find a response assessment by such criteria from the articles, and we determined responder/non-responder using alternative clinical data available. For example, in the datasets from Snyder et al [32]. and from Anagnostou et al [33], where survival outcome was available, after excluding cases who survived less than six months of follow-up, cases with a PFS of 12 months or more were determined to be responders, and others non-responders. For the dataset from Cristescu et al [25], we distinguished between responders and non-responders based on the values listed in the figures and tables in the paper, and confirmed that the annotations were consistent with the results of the other figures. As a result, a total of 973 patients from 13 datasets that met the above criteria and had available response information were included in the analysis (Table S3).

**WES analysis pipeline**

For the raw paired WES data obtained from NBDC, dbGaP and EGA (Table S2, Table S3), somatic mutations were analyzed in the following steps according to the Best Practice Workflows of somatic short variant (SNVs + Indels) discovery published by the Broad institute GATK team [34], where Genome Analysis Toolkit (GATK) v4.0.12 and Picard v2.20.2 were used. (1) BAM files were converted to FASTQ files using Picard SamToFastq. (2) Low-quality reads and presumed adapter sequences were removed using TrimGalore [35]. (3) The trimmed reads were aligned to the human reference genome hg38 obtained from the GATK resource bundle using BWA mem (v0.7.17-r1188) [36]. (4) Data preprocessing including marking duplicates, making the panel of normal samples, and estimating contamination was performed using the Picard and GATK tools. (5) Somatic mutation calling including orientation bias filtering was performed using Mutect2. (6) The following parameters were used to filter out variants with low reliability: annotated as "PASS"; "TLOD" greater than 6.3 and 9 for single nucleotide variants and insertions and deletions, respectively; coverage of altered alleles greater than or equal to 5; altered allele frequency greater than or equal to 2.5%. (7) Variant annotation was performed using Funcotator with default setting using data source of v1.7.20200521s to generate MAF format files.

For the dataset from Hellmann et al [37], only the VCF files after somatic mutation calling were available, so we filtered variants using the same criteria as described above and converted to MAF files for subsequent analysis.

**Validation of the WES analysis pipeline**

As reported by Litchfield et al [38], TMB does not differ significantly depending on the exome capture kits used. For compatibility between TCGA and other datasets, we adopted the Agilent SureSelect Human All Exon V5 capture kit as the common exome regions to be analyzed in our WES analysis pipeline. Comparing the number of missense mutations calculated from cBioPortal [1] with those from our pipeline among the same samples, the Pearson correlation tended to exceed 0.9 in most of the datasets (Figure S19). Similarly, when we compared the number of non-synonymous SNVs in the Cristescu et al paper [25] with those from our pipeline, the Pearson correlation was 0.945 (Figure S19).

**TcellExTRECT score**

The BAM files derived from the tumor samples mapped in the above pipeline were used as input to calculate the TcellExTRECT scores [26] in the same way as the TCGA data analysis.

#### Statistical analyses

Unless otherwise noted, statistical analyses were performed in Python (3.7.4). The Mann–Whitney U test, chi-square test, and Spearman's rank correlation coefficient test were performed using SciPy (1.6.1). Survival analyses including the Kaplan–Meier curve, log-rank test, and Cox proportional hazard regression model were performed using Lifelines (0.25.10) and StatsModels (0.12.2). Machine learning analyses were performed using Scikit-learn (0.24.1). We considered a p-value < 0.05 as being statistically significant. Venn diagrams were depicted using "*VennDiagram*" (1.6.20) package in R. The Jonckheere-Terpstra test was performed using "*clinfun*" (1.0.15) package in R. The Passing-Bablok regression was performed using "*mcr*" (1.2.2) package in R.

#### Data and code availability

Controlled access data used in this study were obtained from dbGaP, EGA, and NBDC with access permissions according to the respective required procedures (Table S2 and S3). The processed data and codes to reproduce the results of this work are available on the GitHub page (https://github.com/shirotak/pancancer_MutSig_ICI). Other codes for preprocessing or restricted-access data are available from the corresponding author upon reasonable request.

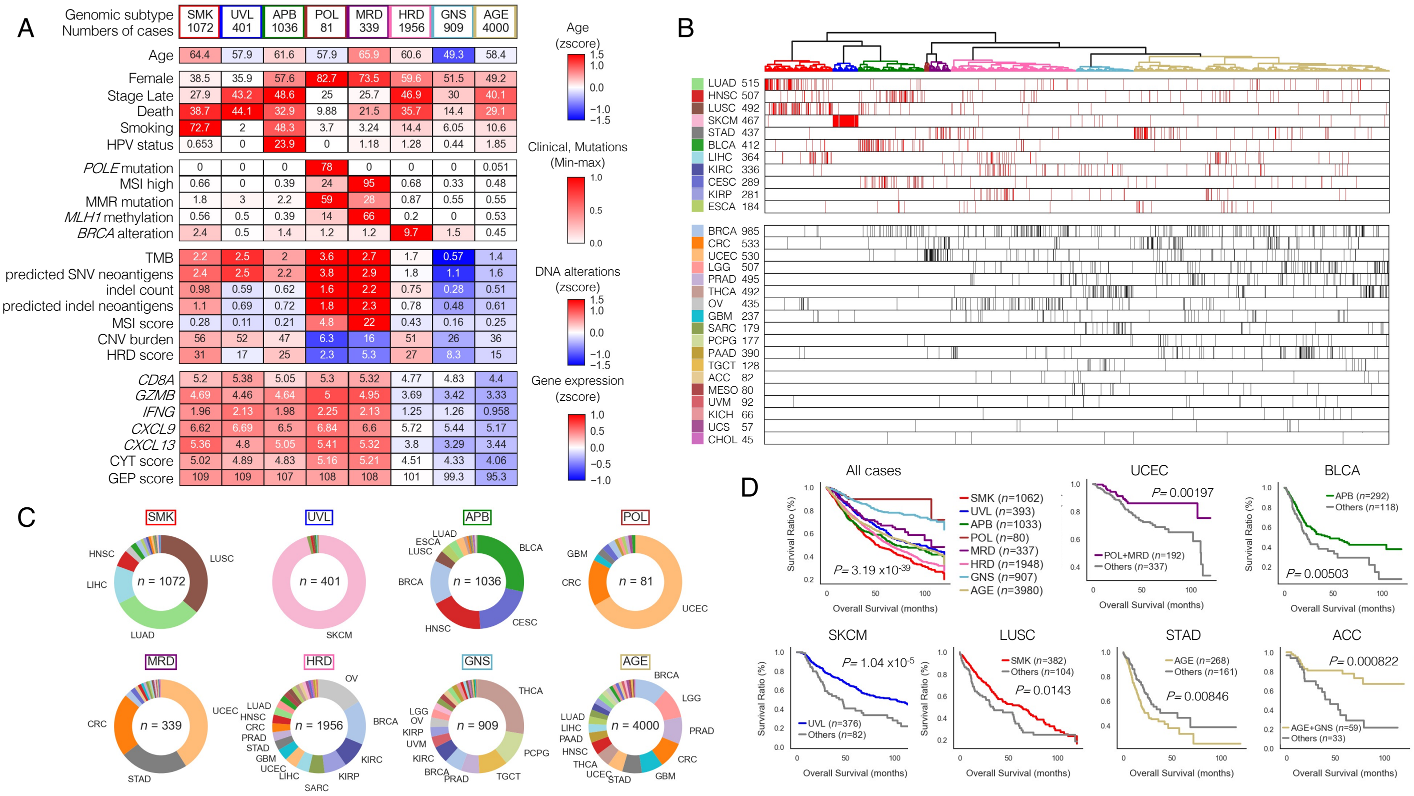

**Figure S1. Characteristics of the eight tumor genomic subtypes derived from TCGA solid tumors (N=9794)**

A) Clinical and genomic information for each subtype, related to Figure1A. The values are either the average of the Z-scores or the frequency for each subtype, and the values are displayed as a heat map. Predicted SNV and indel neoantigens counts by netMHCpan were significantly correlated with TMB and indel counts, respectively. CNV burdens and HRD scores were also correlated with each other.

B) Relationship between the distribution of cancer types based on the hierarchical clustering and the FDA approval for ICI therapy. Each case is represented by a red bar (with FDA approval for ICI) or a black bar (without FDA approval for ICI).

C) Differences in survival outcomes between subtypes. When all cases were compared among the eight subtypes, the prognosis was different. Furthermore, for the cancer types shown here, the prognosis was significantly different by subtype in the analysis of each cancer type. P-value is based on log-rank test.

D) Type of cancer and number of samples per subtype. The color of each cancer type matches the color shown in B.

ACC, Adrenocortical carcinoma; BLCA, Bladder urothelial carcinoma; BRCA, Breast invasive carcinoma; CESC, Cervical squamous cell carcinoma and endocervical adenocarcinoma; CHOL, Cholangiocarcinoma; CRC, Colorectal adenocarcinoma; ESCA, Esophageal carcinoma; GBM, Glioblastoma multiforme; HNSC, Head and neck squamous cell carcinoma; KICH, Kidney chromophobe; KIRC, Kidney renal clear cell carcinoma; KIRP, Kidney renal papillary cell carcinoma; LGG, Brain lower grade glioma; LIHC, Liver hepatocellular carcinoma ; LUAD, Lung adenocarcinoma; LUSC, Lung squamous cell carcinoma; MESO, Mesothelioma; OV, Ovarian serous cystadenocarcinoma; PAAD, Pancreatic adenocarcinoma; PCPG, Pheochromocytoma and paraganglioma; PRAD, Prostate adenocarcinoma; SARC, Sarcoma; SKCM, Skin cutaneous melanoma; STAD, Stomach adenocarcinoma; TGCT, Testicular germ cell tumors; THCA, Thyroid carcinoma; UCEC, Uterine corpus endometrial carcinoma; UCS, Uterine carcinosarcoma; UVM, Uveal melanoma.

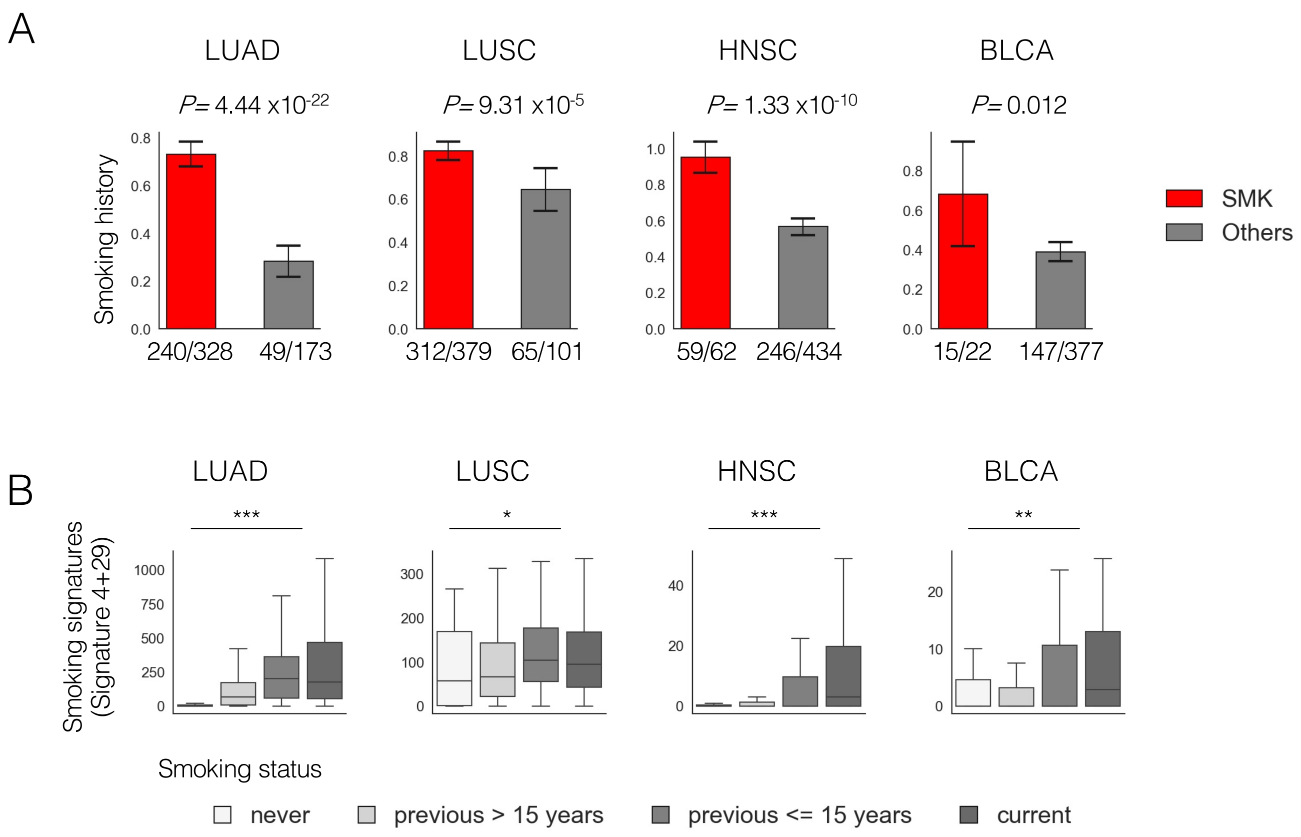


**Figure S2. Features of SMOKING (SMK) subtype (n=1072)**

The association with smoking habits was investigated in LUAD, LUSC, HNSC and BLCA where clinical information on smoking was available.

A) SMK cases had a higher frequency of smoking history (current smoking and smoking within 15 years) than the other groups.

B) The sum of the contributions of signature 4 and 29, which are known to be related to smoking exposure and tobacco chewing habit, were positively correlated with smoking habit. (Jonckheere-Terpstra test, *** $P<1 \times 10^{-4}$, ** $P < 0.01$, *, $P < 0.05$ )

BLCA, Bladder urothelial carcinoma; HNSC, Head and neck squamous cell carcinoma; LUAD, Lung adenocarcinoma; LUSC, Lung squamous cell carcinoma

A

| Cancer type | | Numbers |
|---|---|---|
| SKCM | | 384 |
| Non-SKCM | SARC | 5 |
| | HNSC | 3 |
| | LUSC | 3 |
| | BLCA | 2 |
| | BRCA | 1 |
| | CRC | 1 |
| | GBM | 1 |
| | THCA | 1 |

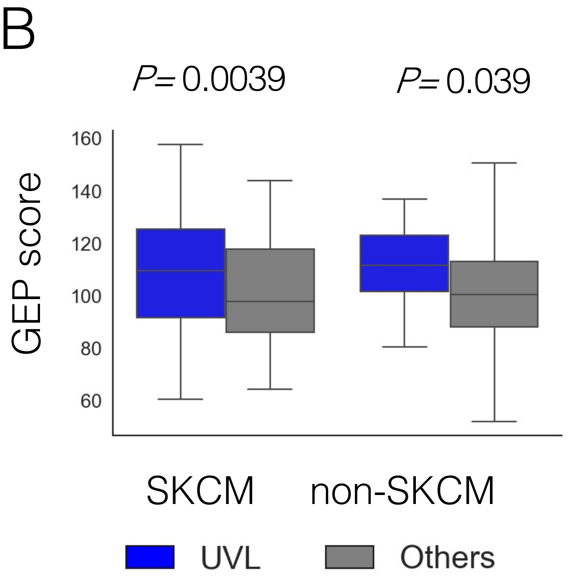


**Figure S3. Features of ULTRAVIOLET LIGHT (UVL) subtype (n=401)**

A) Cancer type of UVL subtype. 95.8% were SKCM.

B) Excluding SKCM, UVL tumors showed higher GEP score than the other subtypes (P= 0.033, Mann-Whitney U test). These results suggest that, although rare in cancers other than SKCM, there are some tumors that exhibit UVL subtype and may have a high tumor immunogenicity.

BLCA, Bladder urothelial carcinoma; BRCA, Breast invasive carcinoma; CRC, Colorectal adenocarcinoma; HNSC, Head and neck squamous cell carcinoma; LUSC, Lung squamous cell carcinoma; SARC, Sarcoma; THCA, Thyroid carcinoma

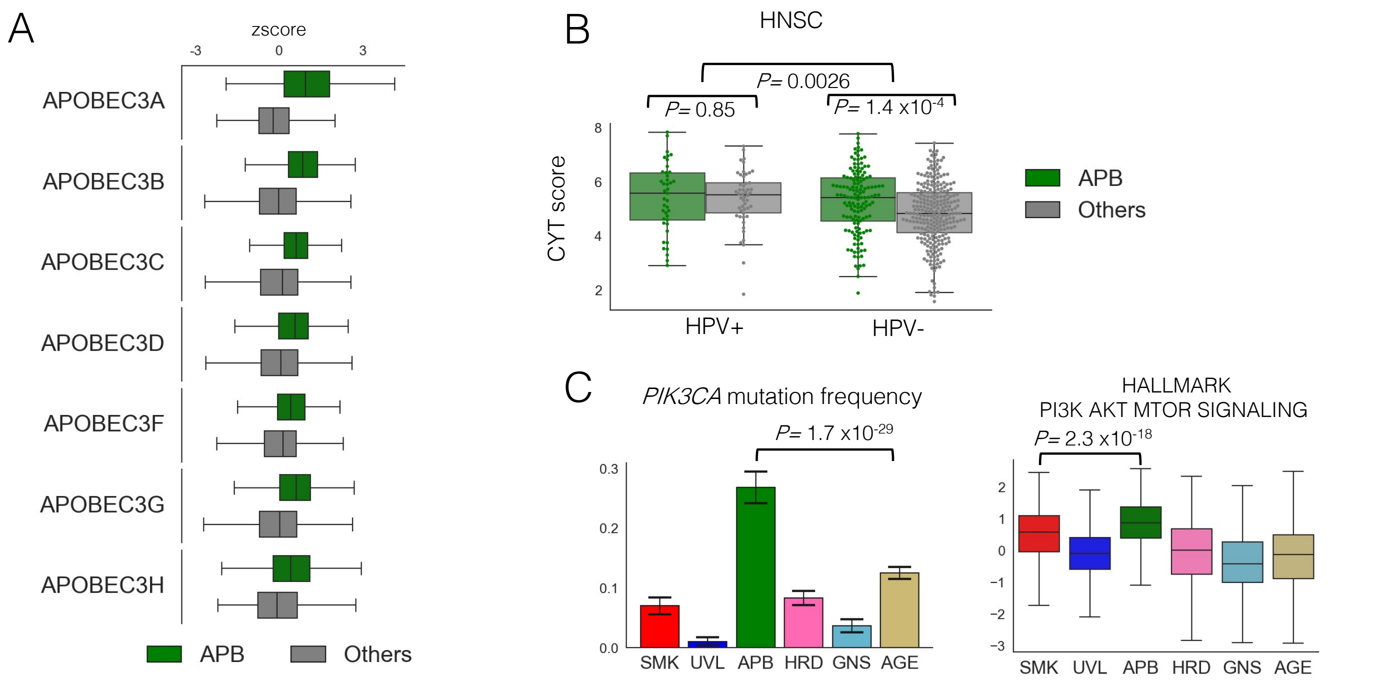


**Figure S4. Features of APOBEC (APB) subtype (n=1036)**

A) The expression of APOBEC3 family genes was significantly higher in APB subtype (all P-values < 2.2 x$10^{-19}$, Mann-Whitney test).

B) HPV infection was associated with higher immune response in TCGA-HNSC (n=507). Besides, even in HPV-negative tumors, APOBEC subtype showed a higher immune response than the other groups.

C) (Left) The *PIK3CA* mutation rate was the highest (26.4%) in the APB subtype excluding hyper mutator subtype (MRD and POL). Chi-square test P=1.7 x$10^{-29}$. (Right), PIK3_AKT_MTOR pathway score was the highest in the APB subtype. Mann-Whitney P=2.3 x$10^{-18}$. These results are consistent with the previously reported association between *PIK3CA* mutation and APOBEC-mediated cytosine deamination [39].

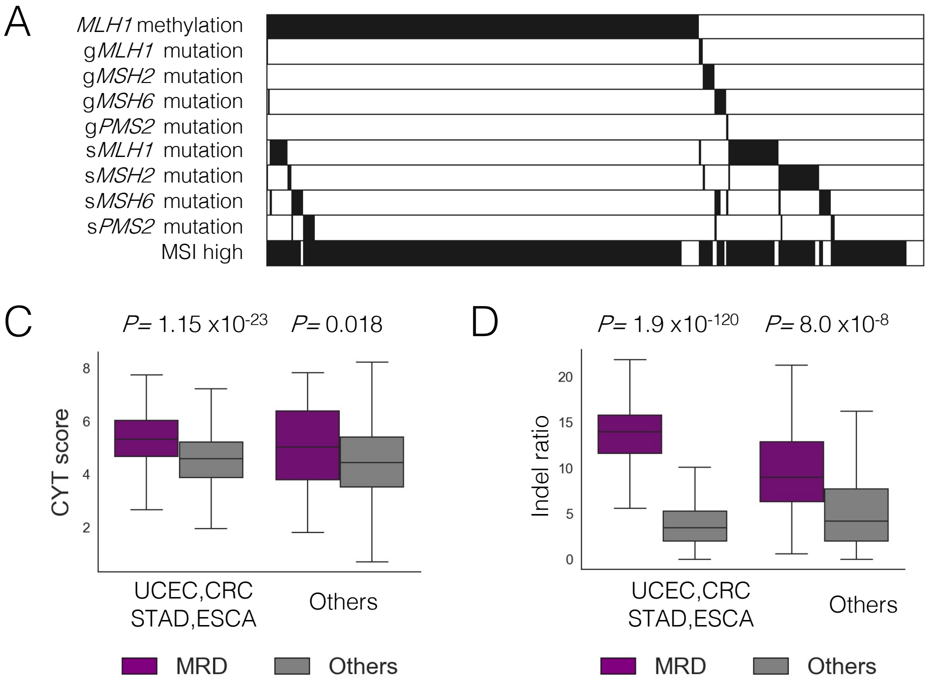


B

| Cancer type | Numbers |
|---|---|
| UCEC | 139 |
| STAD | 80 |
| CRC | 74 |
| ESCA | 2 |
| BRCA | 8 |
| CESC | 6 |
| ACC | 4 |
| HNSC | 4 |
| GBM | 3 |
| SARC | 3 |
| PRAD | 3 |
| LUSC | 2 |
| LIHC | 2 |
| LUAD | 1 |
| LGG | 1 |
| MESO | 1 |
| OV | 1 |
| KIRC | 1 |
| KICH | 1 |
| SKCM | 1 |
| CHOL | 1 |
| UCS | 1 |

**Figure S5. Features of MISMATCH REPAIR DEFICIENCY (MRD) subtype (n=339)**

A) 86.4% had MMR gene alterations (somatic and germline mutation in *MLH1, MSH2, MSH6,* and *PMS2, and MLH1* methylation).

B) Cancer types of MRD subtype.

C) Excluding cancer types with relatively high frequently of MSI-high tumors (UCEC, CRC, STAD, ESCA), immune-related gene expression scores were still higher in MRD subtype than the others (P=0.016, Man-Whitney U test)

D) Excluding cancer types with relatively high frequently of MSI-high tumors (UCEC, CRC, STAD, ESCA), indel ratio was still higher in MRD subtype than the others ($P=4.0 \times 10^{-8}$, Man-Whitney U test)

ACC, Adrenocortical carcinoma; BRCA, Breast invasive carcinoma; CESC, Cervical squamous cell carcinoma and endocervical adenocarcinoma; CHOL, Cholangiocarcinoma; CRC, Colorectal adenocarcinoma; ESCA, Esophageal carcinoma; GBM, Glioblastoma multiforme; HNSC, Head and neck squamous cell carcinoma; KICH, Kidney chromophobe; KIRC, Kidney renal clear cell carcinoma; LGG, Brain lower grade glioma; LIHC, Liver hepatocellular carcinoma ; LUAD, Lung adenocarcinoma; LUSC, Lung squamous cell carcinoma; MESO, Mesothelioma; OV, Ovarian serous cystadenocarcinoma; PRAD, Prostate adenocarcinoma; SARC, Sarcoma; SKCM, Skin cutaneous melanoma; STAD, Stomach adenocarcinoma; UCEC, Uterine corpus endometrial carcinoma; UCS, Uterine carcinosarcoma

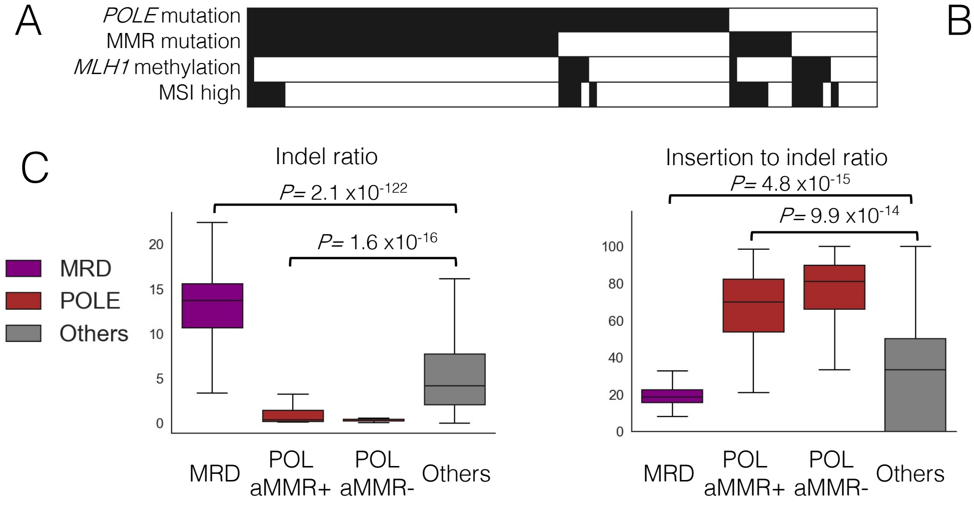


B

| Cancer type | Numbers |
|---|---|
| UCEC | 54 |
| CRC | 13 |
| BRCA | 2 |
| CESC | 2 |
| GBM | 2 |
| STAD | 2 |
| BLCA | 1 |
| ESCA | 1 |
| LGG | 1 |
| PAAD | 1 |
| PRAD | 1 |
| UCS | 1 |

**Figure S6. Features of *POLE* (POL) subtype (n=81)**

A) Somatic *POLE* mutation was observed in 76.5% of POL subtype, and MMR-gene mutation (somatic and germline mutation in *MLH1, MSH2, MSH6,* and *PMS2*) was observed in 59.3% of POL subtype.

B) Cancer types of POL subtype

C) POL subtype, both with and without MMR mutation, had low indel ratio (left) and high insertion to indel ratio (right) than other subtypes, in contrast to MRD subtype. These data indicate that most tumors with concurrent *POLE* and MMR-gene mutations have acquired *POLE* mutation first, as previously reported [40], and thus have the *POLE* mutation-dominant underlying mutational processes.

BLCA, Bladder urothelial carcinoma; BRCA, Breast invasive carcinoma; CESC, Cervical squamous cell carcinoma and endocervical adenocarcinoma; CRC, Colorectal adenocarcinoma; ESCA, Esophageal carcinoma; GBM, Glioblastoma multiforme; LGG, Brain lower grade glioma; PAAD, Pancreatic adenocarcinoma; PRAD, Prostate adenocarcinoma; UCEC, Uterine corpus endometrial carcinoma; UCS, Uterine carcinosarcoma

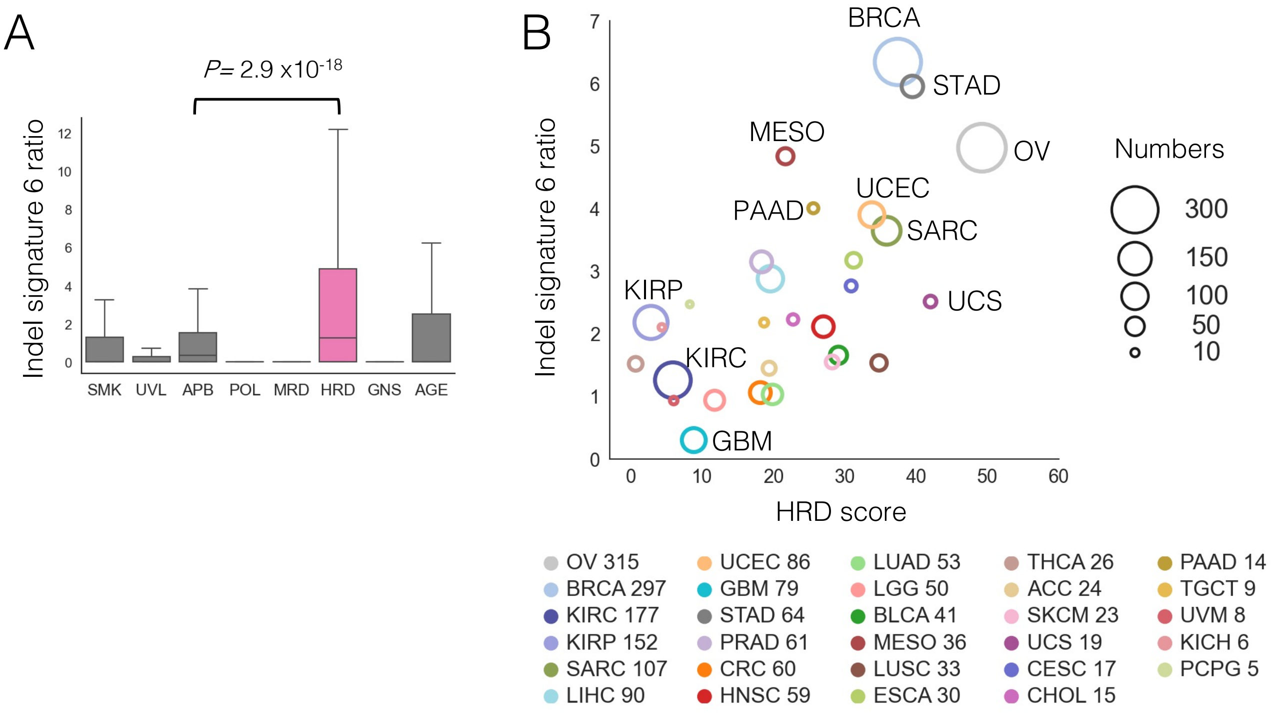


**Figure S7. Features of HOMOLOGOUS RCOMBINATION DEFICIENCY (HRD) subtype (n=1956)**

A) The ratio of indel signature 6, which is related to homologous recombination deficiency, was higher in HRD subtype than in other subtypes.

B) Association between mean HRD scores (x-axis) and indel signature 6 ratios (y-axis) per cancer type calculated in tumors classified into HRD subtype. These two values were simultaneously high in cancer types such as OV, BRCA, SARC, UCEC, and STAD, which are known to include a certain proportion of HRD phenotypes in previous reports [15]. On the other hand, cancer types such as KIRP, KIRC, and GBM showed low values in both, suggesting that the HRD subtype may include tumors which do not harbor HRD phenotype.

ACC, Adrenocortical carcinoma; BLCA, Bladder urothelial carcinoma; BRCA, Breast invasive carcinoma; CESC, Cervical squamous cell carcinoma and endocervical adenocarcinoma; CHOL, Cholangiocarcinoma; CRC, Colorectal adenocarcinoma; ESCA, Esophageal carcinoma; GBM, Glioblastoma multiforme; HNSC, Head and neck squamous cell carcinoma; KICH, Kidney chromophobe; KIRC, Kidney renal clear cell carcinoma; KIRP, Kidney renal papillary cell carcinoma; LGG, Brain lower grade glioma; LIHC, Liver hepatocellular carcinoma ; LUAD, Lung adenocarcinoma; LUSC, Lung squamous cell carcinoma; MESO, Mesothelioma; OV, Ovarian serous cystadenocarcinoma; PAAD, Pancreatic adenocarcinoma; PCPG, Pheochromocytoma and paraganglioma; PRAD, Prostate adenocarcinoma; SARC, Sarcoma; SKCM, Skin cutaneous melanoma; STAD, Stomach adenocarcinoma; TGCT, Testicular germ cell tumors; THCA, Thyroid carcinoma; UCEC, Uterine corpus endometrial carcinoma; UCS, Uterine carcinosarcoma; UVM, Uveal melanoma.

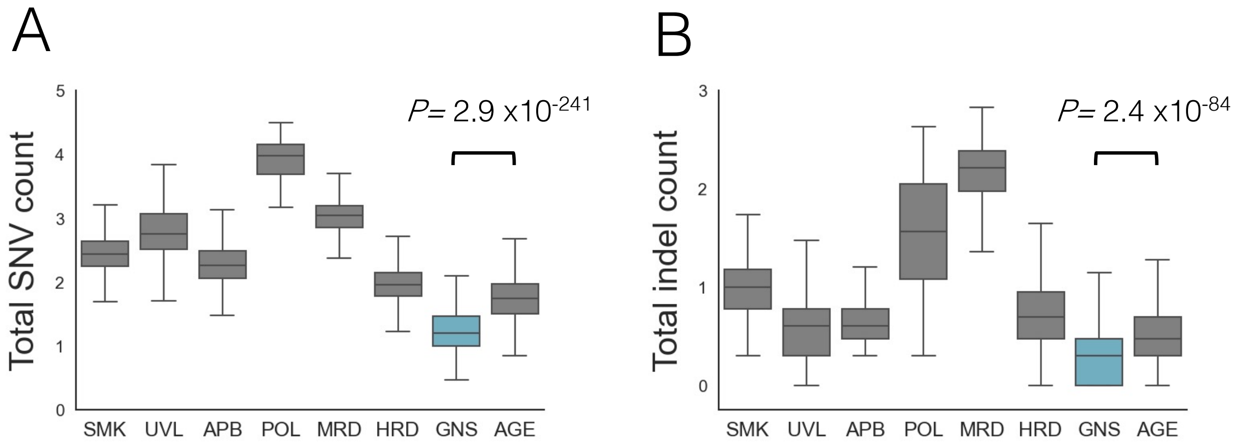


**Figure S8. Features of GENOMICALLY STABLE (GNS) subtype (n=909)**

Not only the total number of SNVs (A), but also that of insertions and deletions (B) were the lowest in GNS among all subtypes.

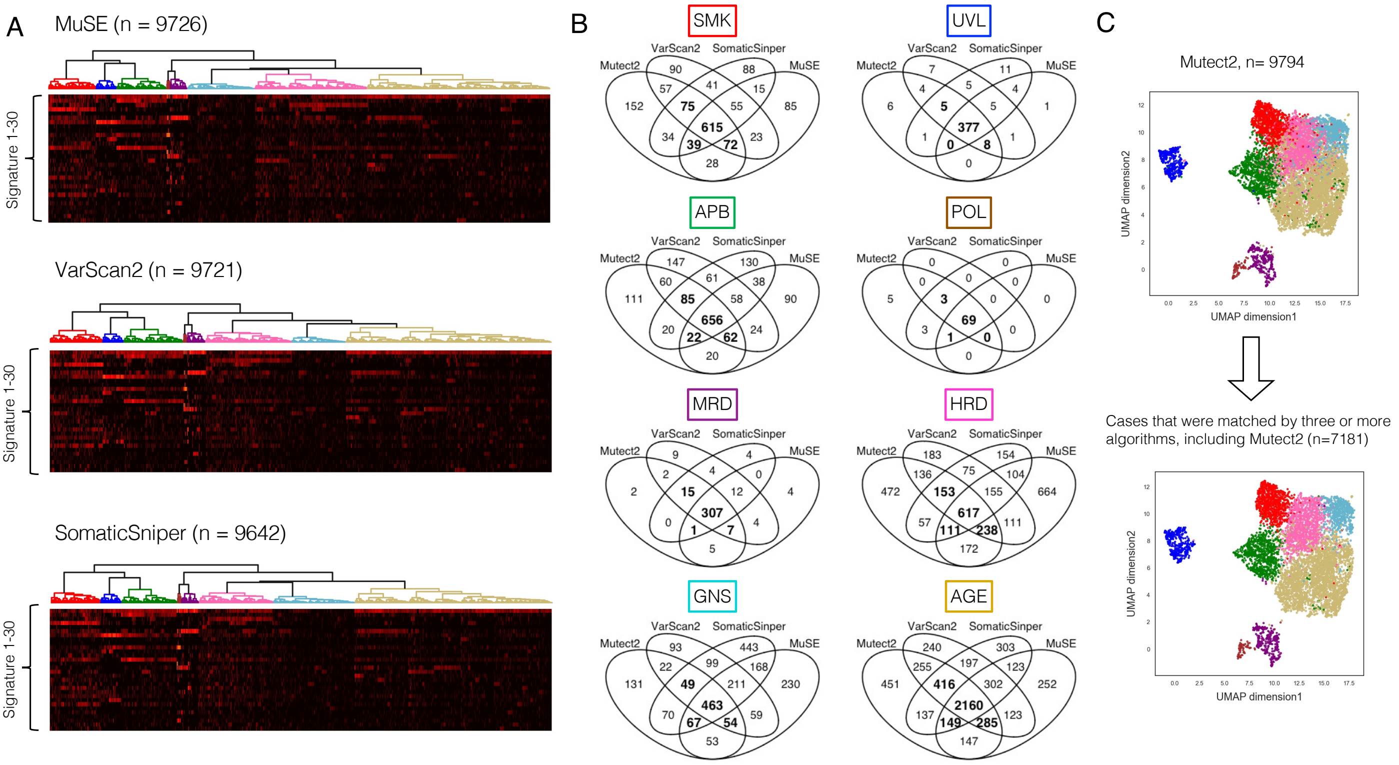


**Figure S9. Consistency of clustering results from different variant callers**

A) The results of hierarchical clustering based on the somatic mutation profiles derived from MuSE (upper), VarScan2 (middle), and SomaticSniper (lower).
Using these methods of annotation, eight genomic subtypes were created, as in the case using Mutect2.

B) Venn diagrams showing the overlapping of results from the four valiant callers per subtype. GNS, HRD, and AGE subtypes showed relatively low consistency. To extract tumors typical for each subtype, cases that were matched by three or more algorithms, including Mutect2, were used for the next analysis (numbers in bold).

C) The UMAP clustering shows that the sample selection described in B reduces the number of cases in the border region of the subtype heatmap.

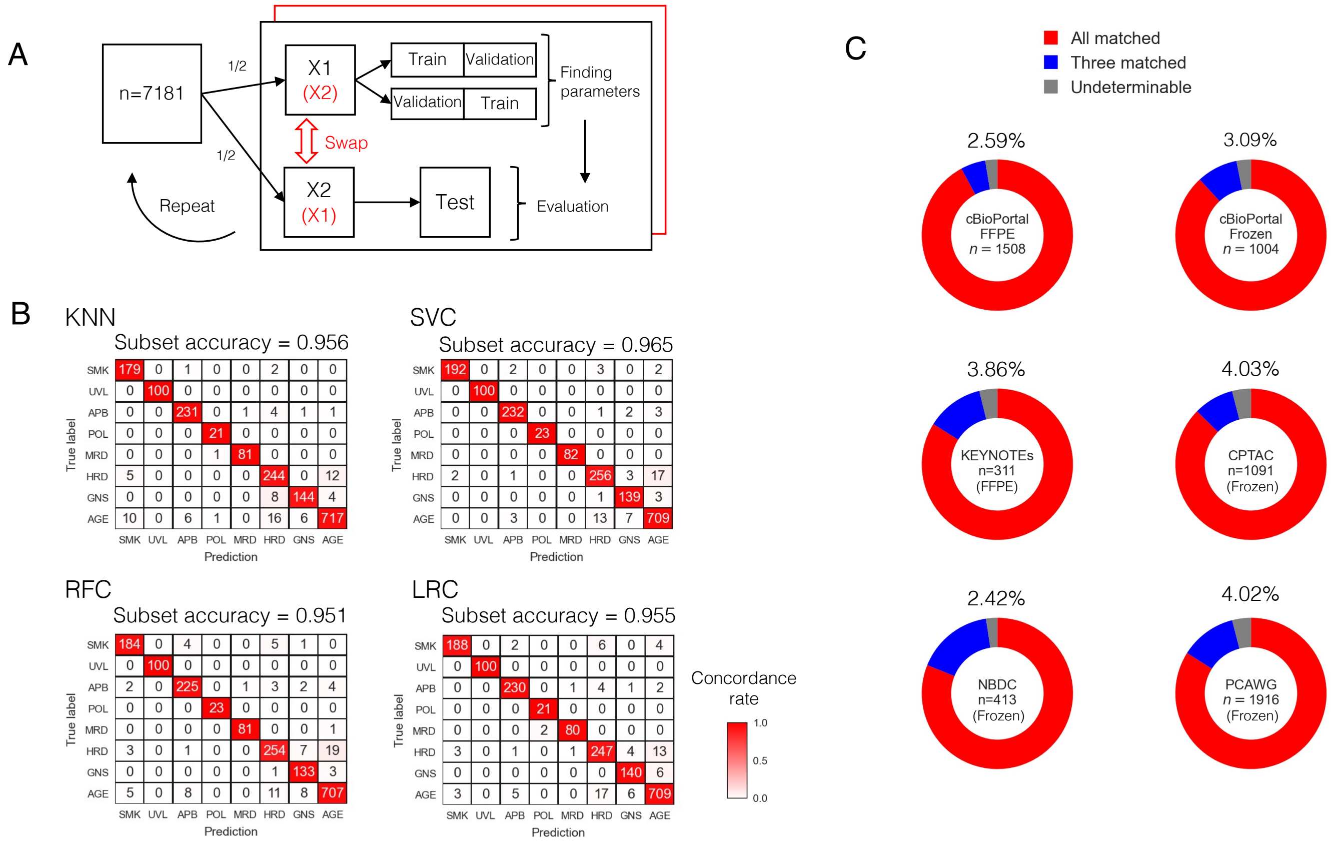


**Figure S10. Developing classifiers through machine learning algorithms**

A) Double cross-validation and hyperparameter tuning. First, the selected 7181 TCGA samples were divided into two parts, X1 and X2. Second, parameters were calculated using X1 by two-fold cross-validation, and those parameters were evaluated using X2 as test data. Third, X1 and X2 were swapped, and the same calculations were performed. These processes were repeated 100-1000 times to determine the optimal parameters.

B) Confusion matrices showing the classification performance on test data for the four classifiers: K-nearest neighbor (KNN), support vector machine (SVC), random forest (RFC), and logistic regression (LRC). Results using 75% of all cases for training and 25% for testing are shown. All showed more then 95% subset accuracy (exact match ratio).

C) Consistency of subtyping results between the four classifiers per data group. When three or more of the four classifier results do not match, the sample is annotated as "undeterminable”. Undeterminable samples were found in approximately 2-4% in the studied data groups, and there was no significant difference in the proportion between FFPE samples and frozen tissue origins or between data groups

A

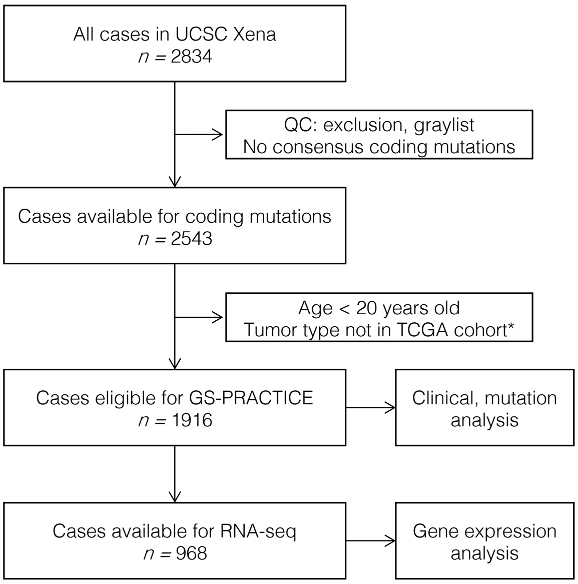


* Excluded: 'CNS-Medullo', 'Lymph-BNHL', 'Lymph-CLL', 'Myeloid-AML', 'Myeloid-AML, Myeloid-MDS', 'Myeloid-AML, Myeloid-MPN', 'Myeloid-MDS', 'Myeloid-MPN', 'Panc-Endocrine, 'Biliary-AdenoCA, Liver-HCC'

B

| TCGA Tumor type | PCAWG Histology abbreviation | Genomic subtype | | | | | | | | |
|---|---|---|---|---|---|---|---|---|---|---|
| | | SMK | UVL | APB | POL | MRD | HRD | GNS | AGE | UND |
| BLCA | Bladder-TCC | 0 | 0 | 13 | 0 | 0 | 2 | 0 | 4 | 4 |
| BRCA | Breast-AdenoCA, LobularCA, DCIS | 0 | 0 | 18 | 0 | 0 | 68 | 11 | 97 | 3 |
| CESC | Cervix-SCC, AdenoCA | 0 | 0 | 10 | 0 | 0 | 0 | 1 | 9 | 0 |
| CHOL | Biliary-AdenoCA | 0 | 0 | 1 | 0 | 1 | 4 | 7 | 18 | 2 |
| CRC | ColoRect-AdenoCA | 1 | 0 | 0 | 7 | 3 | 4 | 0 | 37 | 0 |
| ESCA | Eso-AdenoCA | 0 | 0 | 2 | 0 | 0 | 10 | 1 | 84 | 0 |
| GBM | CNS-GBM | 0 | 0 | 0 | 0 | 0 | 0 | 0 | 38 | 1 |
| HNSC | Head-SCC | 3 | 0 | 17 | 0 | 0 | 5 | 5 | 22 | 3 |
| KICH | Kidney-ChRCC | 0 | 0 | 0 | 0 | 0 | 2 | 16 | 22 | 2 |
| KIRC | Kidney-RCC | 0 | 0 | 0 | 0 | 1 | 92 | 20 | 22 | 8 |
| LGG | CNS-Oligo, PiloAstro | 0 | 0 | 0 | 0 | 0 | 0 | 5 | 20 | 0 |
| LIHC | Liver-HCC | 12 | 0 | 0 | 0 | 2 | 64 | 66 | 139 | 31 |
| LUAD | Lung-AdenoCA | 16 | 0 | 2 | 0 | 0 | 5 | 2 | 8 | 1 |
| LUSC | Lung-SCC | 24 | 0 | 4 | 0 | 0 | 11 | 0 | 1 | 7 |
| OV | Ovary-AdenoCA | 1 | 0 | 0 | 0 | 1 | 78 | 3 | 25 | 2 |
| PAAD | Panc-AdenoCA | 0 | 0 | 0 | 0 | 0 | 24 | 4 | 201 | 2 |
| PRAD | Prost-AdenoCA | 0 | 0 | 0 | 0 | 0 | 14 | 40 | 137 | 5 |
| SKCM | Skin-Melanoma | 0 | 77 | 0 | 0 | 0 | 15 | 1 | 11 | 1 |
| STAD | Stomach-AdenoCA | 0 | 0 | 0 | 0 | 3 | 6 | 4 | 49 | 2 |
| THCA | Thy-AdenoCA | 0 | 0 | 1 | 0 | 0 | 1 | 19 | 21 | 3 |
| UCEC | Uterus-AdenoCA | 0 | 0 | 4 | 1 | 4 | 9 | 0 | 26 | 0 |
| Total | | 57 | 77 | 72 | 8 | 15 | 414 | 205 | 991 | 77 |

**Figure S11. Tumor subtyping by GS-PRACTICE in the PCAWG datasets**

**A)** Sample selection.

From a total of 2834 cases, those included in the whilelist, with consensus coding mutations available, aged 20 years or older, and with tumor types included in TCGA studies were analyzed (n = 1916).

**B)** Distribution of the samples by tumor genomic subtype and by cancer type

PCAWG, Pan-Cancer Analysis of Whole Genomes consortium

A

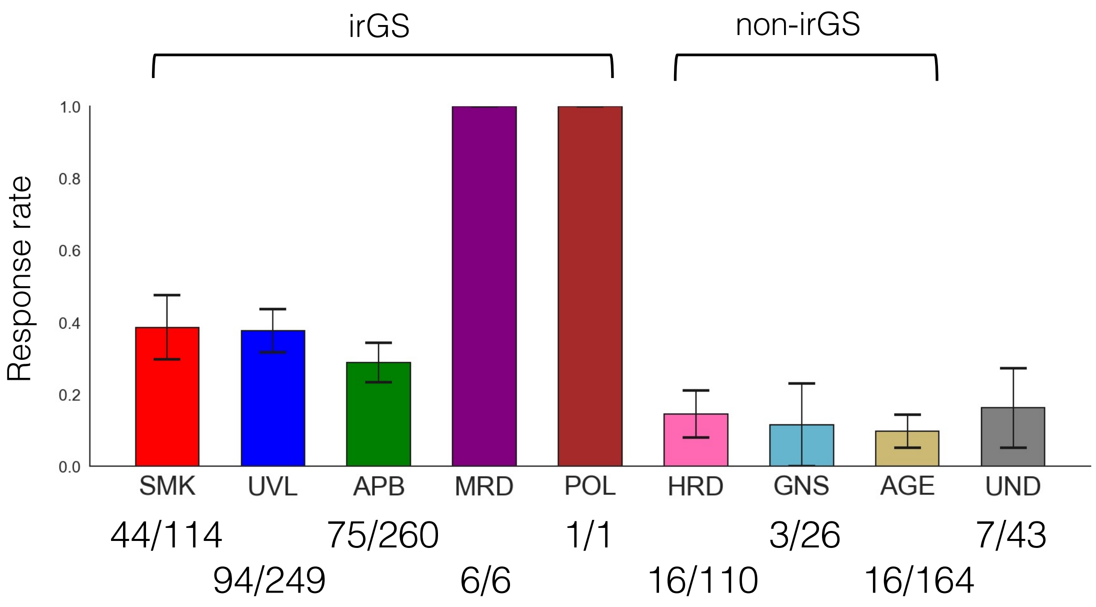


B

| | Genomic subtype | | | | | | | | |
|---|---|---|---|---|---|---|---|---|---|
| Cancer type | SMK | UVL | APB | MRD | POL | HRD | GNS | AGE | UND |
| Melanoma | 2 | 247 | 0 | 0 | 0 | 33 | 12 | 24 | 17 |
| Head and neck | 14 | 2 | 38 | 0 | 0 | 15 | 0 | 43 | 6 |
| Lung | 89 | 0 | 18 | 0 | 0 | 18 | 6 | 26 | 5 |
| Bladder | 8 | 0 | 191 | 4 | 0 | 17 | 0 | 31 | 12 |
| Others | 1 | 0 | 13 | 2 | 1 | 27 | 8 | 40 | 3 |

**Figure S12. Relationship between tumor genomic subtype, response rate and cancer type in the ICI-treated cohort**

A) ICI response rate for each tumor genomic subtype. Subtypes included in irGS (SMK, UVL, APB, MRD, POL) showed a higher response rate than subtypes included in non-irGS (HRD, GNS, AGE).

B) Distribution of the samples by tumor genomic subtype and by cancer type.

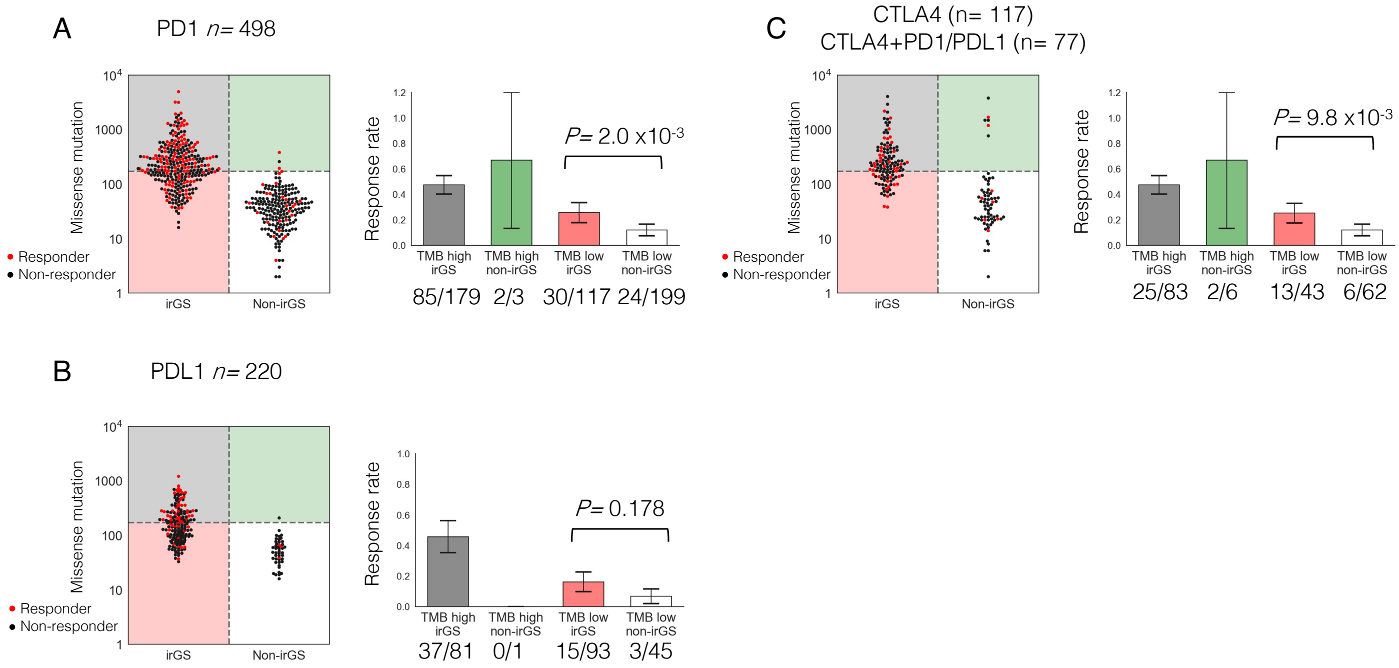


**Figure S13. Studies examined by type of drug**

A) Study in the cases treated with anti-PD1 antibody.

B) Study in the cases treated with anti-PDL1 antibody.

C) Study in the cases treated with anti-CTLA antibody or anti-CTLA antibody plus other ICIs.

In all studies, ICI response rate tended to be higher in irGS compared to non-irGS within TMB low cases.

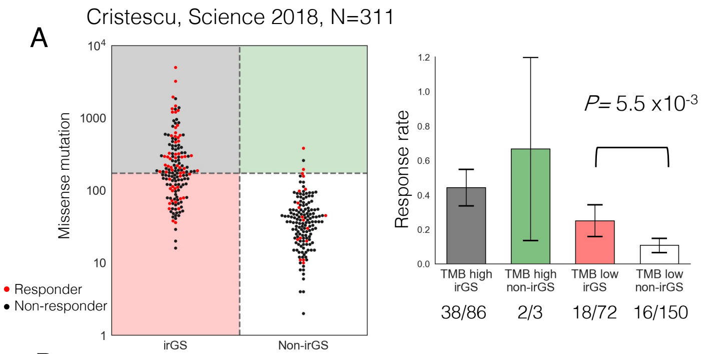


B

| All cases (N=311) | | | | Univariate | | | Multivariate | | |
|---|---|---|---|---|---|---|---|---|---|
| Variables | | No. | Response rate (%) | OR | 95%CI | P-val | OR | 95%CI | P-val |
| irGS | No | 153 | 11.8 | | | | | | |
| | Yes | 158 | 35.4 | 4.12 | (2.28 - 7.43) | $2.6 \times 10^{-6}$ | 2.16 | (1.01 - 4.61) | 0.046 |
| TMB | low | 222 | 15.3 | | | | | | |
| | high | 89 | 44.9 | 4.51 | (2.59 - 7.86) | $1 \times 10^{-7}$ | 2.10 | (1.03 - 4.30) | 0.042 |
| Cancer type | Melanoma | 89 | 42.7 | | | | | | |
| | Head neck | 106 | 18.9 | 0.31 | (0.16 - 0.59) | $3.8 \times 10^{-4}$ | 0.52 | (0.26 - 1.07) | 0.075 |
| | Bladder | 17 | 17.6 | 0.29 | (0.08 - 1.07) | 0.063 | 0.32 | (0.08 - 1.24) | 0.10 |
| | Others | 99 | 13.1 | 0.20 | (0.10 - 0.42) | $1.4 \times 10^{-5}$ | 0.47 | (0.20 - 1.10) | 0.081 |
| Sex | Female | 117 | 18.8 | | | | | | |
| | Male | 194 | 26.8 | 1.58 | (0.90 - 2.77) | 0.11 | | | |
| Age | < 62 | 151 | 25.6 | | | | | | |
| | >= 62 | 160 | 21.9 | 0.82 | (0.48 - 1.37) | 0.44 | | | |

C

| | Genomic subtype | | | | | | | | Total |
|---|---|---|---|---|---|---|---|---|---|
| Cancer type | SMK | UVL | APB | MRD | HRD | GNS | AGE | UND | |
| Head and neck SCC | 13 | 2 | 35 | 0 | 14 | 0 | 36 | 6 | 106 |
| Melanoma | 0 | 72 | 0 | 0 | 10 | 0 | 4 | 3 | 89 |
| Urothelial | 1 | 0 | 14 | 0 | 0 | 0 | 2 | 0 | 17 |
| Breast | 0 | 0 | 2 | 0 | 6 | 0 | 4 | 2 | 14 |
| Gastric | 0 | 0 | 0 | 1 | 1 | 0 | 8 | 0 | 10 |
| Salivary Gland | 0 | 0 | 1 | 0 | 7 | 0 | 2 | 0 | 10 |
| Thyroid Cancer | 0 | 0 | 0 | 0 | 3 | 2 | 4 | 0 | 9 |
| Analcanal SCC | 0 | 0 | 6 | 0 | 0 | 0 | 2 | 0 | 8 |
| Mesothelioma | 0 | 0 | 0 | 0 | 2 | 3 | 1 | 1 | 7 |
| Biliary Tract | 0 | 0 | 0 | 1 | 2 | 1 | 2 | 0 | 6 |
| Carcinoid | 1 | 0 | 0 | 0 | 2 | 0 | 3 | 0 | 6 |
| Colorectal Adeno | 0 | 0 | 0 | 0 | 1 | 0 | 4 | 0 | 5 |
| Small cell lung cancer | 3 | 0 | 0 | 0 | 0 | 0 | 1 | 0 | 4 |
| Endometrial | 0 | 0 | 0 | 0 | 0 | 0 | 3 | 0 | 4 |
| Vulvar SCC | 0 | 0 | 2 | 0 | 0 | 0 | 1 | 0 | 3 |
| ProstateAdeno | 0 | 0 | 0 | 0 | 1 | 1 | 1 | 0 | 3 |
| Esophageal SCC/Adeno | 0 | 0 | 1 | 0 | 1 | 0 | 1 | 0 | 3 |
| Cervical SCC | 0 | 0 | 1 | 0 | 0 | 1 | 1 | 0 | 3 |
| Pancreas Adeno | 0 | 0 | 0 | 0 | 0 | 0 | 2 | 0 | 2 |
| Leiomyosarcoma | 0 | 0 | 0 | 0 | 1 | 0 | 1 | 0 | 2 |

**Figure S14. Study in the dataset from the KEYNOTE trials, which were prospective cohort studies of patients treated with solely pembrolizumab (n=311)**

A) Association between TMB and ICI response per sample divided by irGS status (left) and comparison of ICI response rate in the four groups stratified by irGS and TMB status (right). irGS showed a significantly higher response rate than non-irGS within the sample classified as TMB low.

B) Univariate and multivariate logistic regression analysis for ICI response. irGS status was significantly associated with the ICI response after adjusting by TMB status and cancer type.

C) Distribution of the samples by tumor genomic subtype and by cancer type. Although the KEYNOTE trials excluded patients with clinically diagnosed MSI high tumors at enrollment, two tumors from the cohort (one each with gastric cancer and biliary tract cancer) were classified as MRD subtype, and both of them responded to ICI.

irGS, immune-reactive genomic subtype; TMB, Tumor mutational burden; OR, Odds ratio; CI, confidence interval; SCC, squamous cell carcinoma

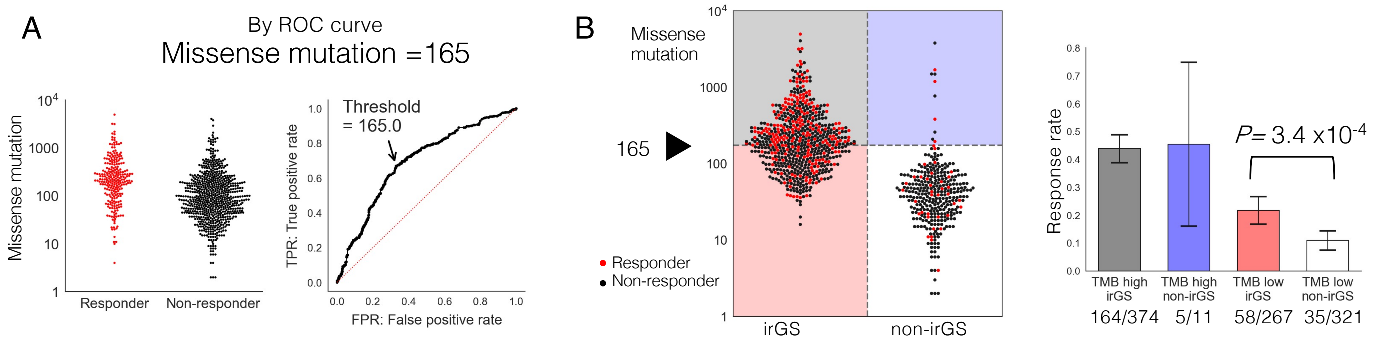


C

| All cases (N=973) | | | | Univariate | | | Multivariate (TMB binary) | | | Multivariate (TMB log10) | | |
|---|---|---|---|---|---|---|---|---|---|---|---|---|
| Variables | | No. | Response rate (%) | OR | 95%CI | p-value | OR | 95%CI | p-value | OR | 95%CI | p-value |
| irGS | No | 332 | 12.0 | | | | | | | | | |
| | Yes | 641 | 34.6 | 3.87 | (2.68 - 5.59) | 6.1 x$10^{-13}$ | 2.02 | (1.29 - 3.17) | 2.2 x$10^{-3}$ | 1.90 | (1.20 - 3.03) | 6.5 x$10^{-3}$ |
| TMB | low | 588 | 15.8 | | | | | | | | | |
| | high | 385 | 43.9 | 4.16 | (3.09 - 5.62) | 9.5 x$10^{-21}$ | 2.95 | (2.06 - 4.22) | 3.2 x$10^{-9}$ | | | |
| TMB log10 | | 973 | | 3.92 | (2.87 - 5.37) | 1.2 x$10^{-17}$ | | | | 2.77 | (1.86 - 4.11) | 5.1 x$10^{-7}$ |
| Cancer Type | Melanoma | 335 | 32.2 | | | | | | | | | |
| | Head neck | 118 | 18.6 | 0.48 | (0.29 - 0.81) | 5.6 x$10^{-3}$ | 0.84 | (0.48 - 1.47) | 0.55 | 0.84 | (0.48 - 1.45) | 0.53 |
| | Lung | 162 | 30.2 | 0.91 | (0.61 - 1.37) | 0.64 | 1.22 | (0.79 - 1.88) | 0.37 | 1.21 | (0.78 - 1.86) | 0.40 |
| | Bladder | 263 | 26.6 | 0.76 | (0.53 - 1.09) | 0.14 | 0.91 | (0.62 - 1.33) | 0.62 | 0.90 | (0.61 - 1.31) | 0.57 |
| | Others | 95 | 13.7 | 0.33 | (0.18 - 0.62) | 6.1 x$10^{-4}$ | 0.90 | (0.45 - 1.79) | 0.75 | 0.89 | (0.44 - 1.79) | 0.75 |
| ICI agent | Double | 77 | 31.2 | | | | | | | | | |
| | Single | 896 | 26.6 | 0.80 | (0.48 - 1.32) | 0.38 | | | | | | |
| Sex | Female | 342 | 24.3 | | | | | | | | | |
| | Male | 631 | 28.4 | 1.24 | (0.91 - 1.67) | 0.17 | | | | | | |
| Age | < 62 | 334 | 25.4 | | | | | | | | | |
| | >= 62 | 339 | 26.8 | 1.08 | (0.76 - 1.52) | 0.68 | | | | | | |

**Figure S15. Studies using the cohorts' optimal TMB cutoff or logarithmic TMB as a continuous value**

A) The cohort's optimal TMB cutoff determined by the ROC curve and the Youden index for objective responses in the whole cohort (N=973) was 165 missense mutations, which was close to 173, the value calculated Figure3B.

B) Association between TMB and ICI response per sample divided by irGS status (left) and comparison of ICI response rate in the four groups stratified by irGS and TMB status (right). irGS showed a significantly higher response rate than non-irGS within the sample classified as TMB low.

C) Univariate and multivariate logistic regression analysis for ICI response in the validation cohort (N=973). irGS status was significantly associated with the ICI response after adjusting by TMB status, either as a binary or continuous value, in addition to cancer type.

irGS, immune-reactive genomic subtype; TMB, Tumor mutational burden; OR, Odds ratio; CI, confidence interval

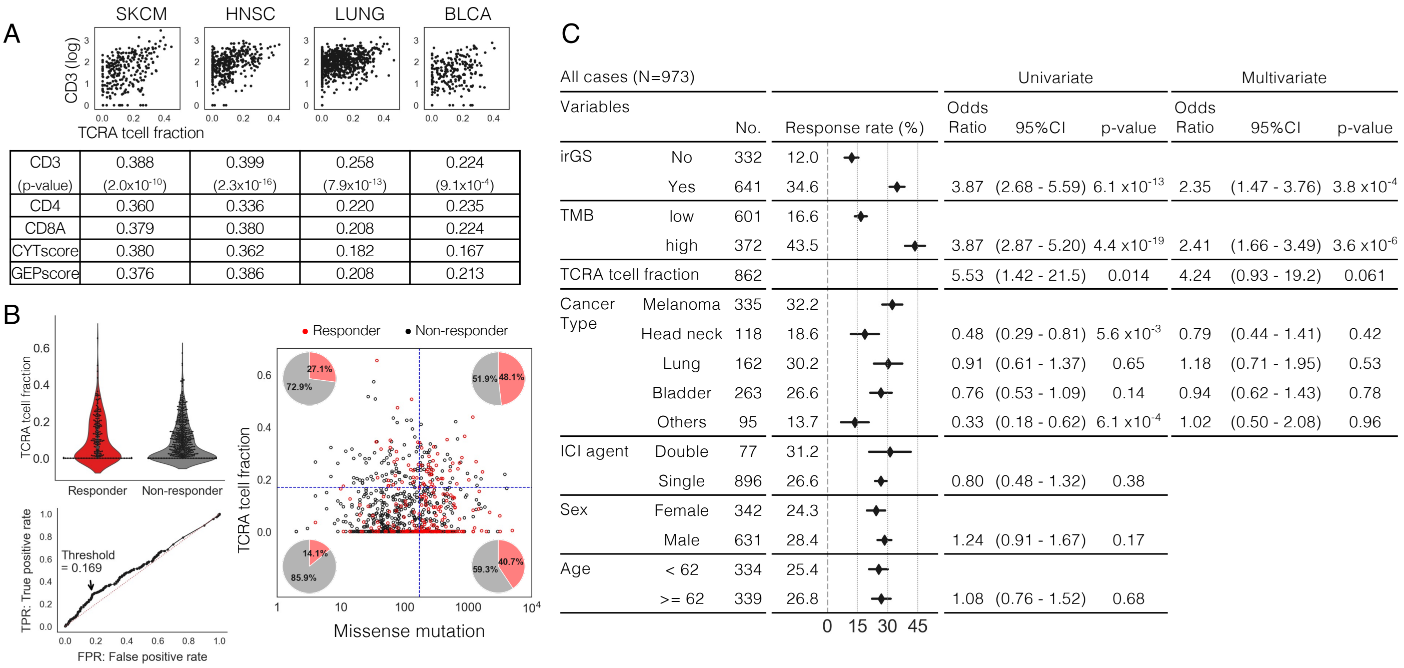


| | SKCM | HNSC | LUNG | BLCA |
|---|---|---|---|---|
| CD3 (p-value) | 0.388 ($2.0x10^{-10}$) | 0.399 ($2.3x10^{-16}$) | 0.258 ($7.9x10^{-13}$) | 0.224 ($9.1x10^{-4}$) |
| CD4 | 0.360 | 0.336 | 0.220 | 0.235 |
| CD8A | 0.379 | 0.380 | 0.208 | 0.224 |
| CYTscore | 0.380 | 0.362 | 0.182 | 0.167 |
| GEPscore | 0.376 | 0.386 | 0.208 | 0.213 |

| All cases (N=973) | | | | Univariate | | | Multivariate | | |
|---|---|---|---|---|---|---|---|---|---|
| Variables | | No. | Response rate (%) | Odds Ratio | 95%CI | p-value | Odds Ratio | 95%CI | p-value |
| irGS | No | 332 | 12.0 | | | | | | |
| | Yes | 641 | 34.6 | 3.87 | (2.68 - 5.59) | $6.1 x10^{-13}$ | 2.35 | (1.47 - 3.76) | $3.8 x10^{-4}$ |
| TMB | low | 601 | 16.6 | | | | | | |
| | high | 372 | 43.5 | 3.87 | (2.87 - 5.20) | $4.4 x10^{-19}$ | 2.41 | (1.66 - 3.49) | $3.6 x10^{-6}$ |
| TCRA tcell fraction | | 862 | | 5.53 | (1.42 - 21.5) | 0.014 | 4.24 | (0.93 - 19.2) | 0.061 |
| Cancer Type | Melanoma | 335 | 32.2 | | | | | | |
| | Head neck | 118 | 18.6 | 0.48 | (0.29 - 0.81) | $5.6 x10^{-3}$ | 0.79 | (0.44 - 1.41) | 0.42 |
| | Lung | 162 | 30.2 | 0.91 | (0.61 - 1.37) | 0.65 | 1.18 | (0.71 - 1.95) | 0.53 |
| | Bladder | 263 | 26.6 | 0.76 | (0.53 - 1.09) | 0.14 | 0.94 | (0.62 - 1.43) | 0.78 |
| | Others | 95 | 13.7 | 0.33 | (0.18 - 0.62) | $6.1 x10^{-4}$ | 1.02 | (0.50 - 2.08) | 0.96 |
| ICI agent | Double | 77 | 31.2 | | | | | | |
| | Single | 896 | 26.6 | 0.80 | (0.48 - 1.32) | 0.38 | | | |
| Sex | Female | 342 | 24.3 | | | | | | |
| | Male | 631 | 28.4 | 1.24 | (0.91 - 1.67) | 0.17 | | | |
| Age | < 62 | 334 | 25.4 | | | | | | |
| | >= 62 | 339 | 26.8 | 1.08 | (0.76 - 1.52) | 0.68 | | | |

**FigureS16. Study using TcellExTRECT score**

A) Spearman correlations between the TCRA tcell fraction score, calculated by TcellExTRECT [26], versus T-cell related gene expression scores for each cancer type in TCGA data. CD3 expression levels were calculated as the geometric mean of CD3D, CD3E, and CD3G expression levels. These data indicate that the score can certainly reflect the immune cell infiltration in the tumor.

B) Associations between ICI response, TCRA tcell fraction, and tumor mutational burden in the validation cohort. The TCRA tcell fraction score was successfully calculated in 862 samples. We calculated the optimal cutoff of the score for ICI response (=0.169), and validated that the score correlated with ICI response independently of tumor mutational burden.

C) Univariate and multivariate logistic regression analysis for ICI response in the validation cohort with the TCRA tcell fraction score as a covariate. irGS status was significantly associated with the ICI response after adjusting by TMB status, TCRA tcell fraction, and cancer type.

irGS, immune-reactive genomic subtype; TMB, Tumor mutational burden; OR, Odds ratio; CI, confidence interval

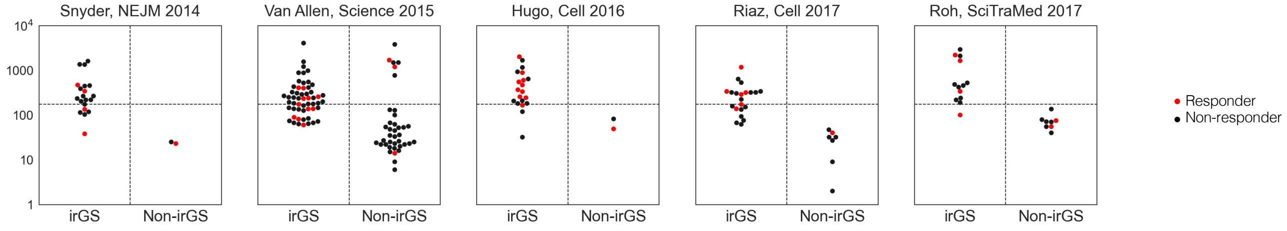


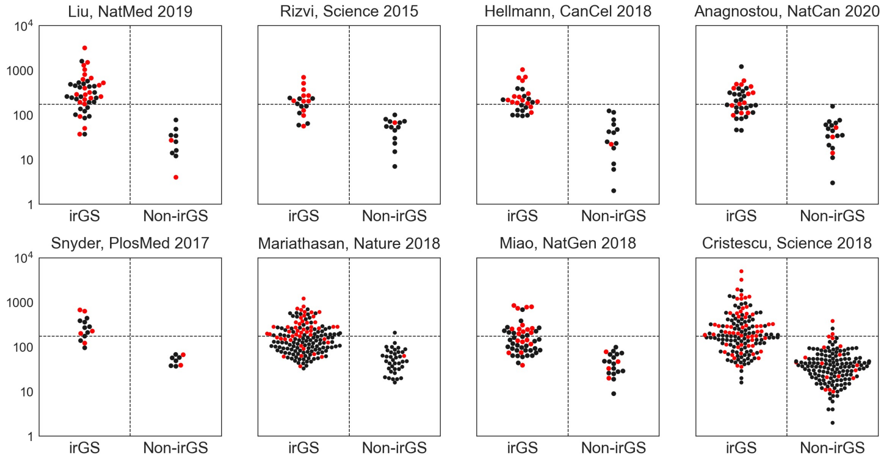


| | Genomic subtype | | | | | | | | |
|---|---|---|---|---|---|---|---|---|---|
| Reference | SMK | UVL | APB | MRD | POL | HRD | GNS | AGE | UND |
| Snyder, NEJM 2014 | 0 | 20 | 0 | 0 | 0 | 1 | 0 | 1 | 0 |
| Van Allen, Science 2015 | 0 | 54 | 0 | 0 | 0 | 9 | 7 | 10 | 12 |
| Hugo, Cell 2016 | 0 | 20 | 0 | 0 | 0 | 1 | 0 | 0 | 1 |
| Riaz, Cell 2017 | 1 | 20 | 0 | 0 | 0 | 3 | 1 | 3 | 0 |
| Roh, SciTraMed 2017 | 1 | 11 | 0 | 0 | 0 | 4 | 0 | 4 | 1 |
| Liu, NatMed 2019 | 0 | 45 | 0 | 0 | 0 | 4 | 4 | 2 | 0 |
| Rizvi, Science 2015 | 17 | 0 | 2 | 0 | 0 | 6 | 1 | 5 | 3 |
| Hellmann, CanCel 2018 | 27 | 0 | 2 | 0 | 0 | 2 | 4 | 8 | 0 |
| Anagnostou, NatCan 2020 | 31 | 0 | 5 | 0 | 0 | 6 | 1 | 10 | 1 |
| Snyder, PlosMed 2017 | 0 | 0 | 12 | 1 | 0 | 1 | 0 | 7 | 1 |
| Mariathasan, Nature 2018 | 6 | 0 | 144 | 3 | 0 | 15 | 0 | 20 | 10 |
| Miao, NatGen 2018 | 13 | 5 | 33 | 0 | 0 | 7 | 0 | 11 | 2 |
| Cristescu, Science 2018 | 18 | 74 | 62 | 2 | 1 | 51 | 8 | 83 | 12 |

**Figure S17. Association between TMB and ICI response divided by irGS status per dataset**

The right table indicates the distribution of samples for each subtype per dataset. See also Table S3.

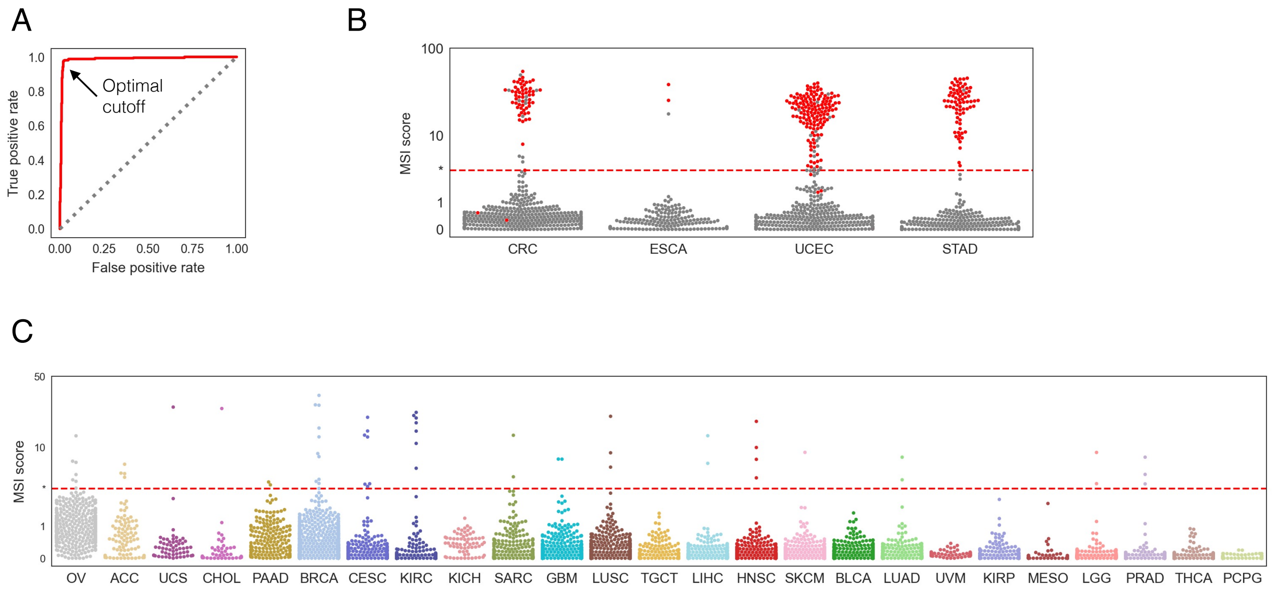


**Figure S18. Determination of MSI-high cases using MSIsensor**

A,B) Optimal cutoff of MSI score to discriminate cases with MSI-high annotations was calculated using ROC curve and Youden index from the datasets of CRC, ESCA, STAD and UCEC.

C) The relationship between MSI score and the cutoff value in cancer types other than the four above. Cases exceeding the cutoff were annotated as MSI high.

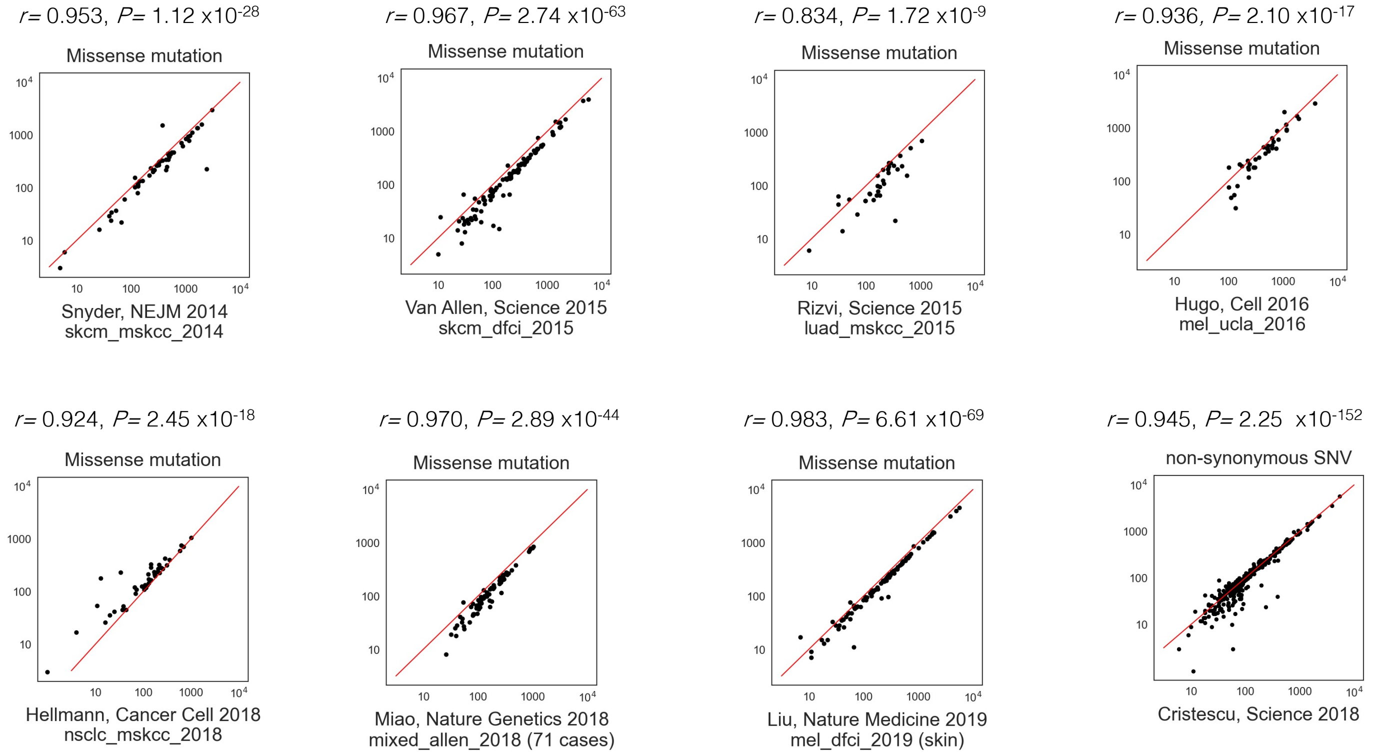


**Figure S19. Comparison of the number of missense mutations or non-synonymous SNVs from our WES pipeline and previously published data**

The red line represents a straight line with slope 1 reaching the zero point. The r value and p value at the top of each panel were calculated using the Pearson's correlation. Most of the datasets have Pearson correlations greater than 0.9.
